# Viral and immune dynamics of HPV genital infections in young women

**DOI:** 10.1101/2023.05.11.23289843

**Authors:** Nicolas Tessandier, Baptiste Elie, Vanina Boué, Christian Selinger, Massilva Rahmoun, Claire Bernat, Sophie Grasset, Soraya Groc, Anne-Sophie Bedin, Thomas Beneteau, Marine Bonneau, Christelle Graf, Nathalie Jacobs, Tsukushi Kamiya, Marion Kerioui, Julie Lajoie, Imène Melki, Jean-Luc Prétet, Bastien Reyné, Géraldine Schlecht-Louf, Mircea T. Sofonea, Olivier Supplisson, Vincent Foulongne, Jérémie Guedj, Christophe Hirtz, Marie-Christine Picot, Jacques Reynes, Vincent Tribout, Édouard Tuaillon, Tim Waterboer, Michel Segondy, Ignacio G Bravo, Nathalie Boulle, Carmen Lia Murall, Samuel Alizon

**Author notes:** equal contribution (Nicolas Tessandier, Baptiste Elie).

## Abstract

Human papillomavirus (HPV) infections drive one in twenty new cancer cases. Despite the potential for improving treatment, screening, and vaccination strategies, little is known as to why most HPV infections clear spontaneously within two years. To untangle the dynamics of these non-persisting infections, we performed a combined quantitative analysis of virological, immunological, and clinical data from an original longitudinal cohort of 189 women with high temporal resolution. We find that HPV viral load reaches a plateau within two months, and clears within a median time of 14 months. Furthermore, we identify immune correlates associated with infection clearance, especially TCR-gamma-delta cells. Our results open new perspectives for understanding the frontier between acute and chronic infections and for controlling HPVassociated diseases.

---

Human papillomaviruses (HPVs) cause nearly all cervical cancers, the majority of many anogenital cancers, and a significant fraction of oropharyngeal cancers (*1*). Women and low-medium income countries are the most affected, with respectively 90% and 65% of the 630,000 HPV-induced cancers reported in 2012 worldwide (*1*). This burden, further exacerbated by millions of cases of anogenital warts (*2*), stems from the fact that HPVs are among the most prevalent sexually-transmitted infections (STIs), with a high transmission risk per sexual contact (*3*). Fortunately, more than 90% of these infections do not persist for more than two years in young adults (*4*–*7*). The factors driving infection clearance are poorly known and could involve the adaptive and the innate immune response (*8*), but also random events occurring during cell division (*9, 10*). Over the last two decades, safe and efficient vaccines have been developed that target the most oncogenic genotypes (especially HPV16 and HPV18), as well as genotypes causing genital warts (HPV6 and HPV11) (*11, 12*). Notwithstanding, chronic infections by HPVs will remain a major public health issue for at least one generation because of low vaccine coverage in many countries and decreased vaccine efficacy if administered after exposition (*13*). Non-persisting, or ‘acute’, HPV infections are often asymptomatic and benign but raise important challenges (*14*). First, the quality of screening policies relies on the description of the natural history of the infection (*15*). Furthermore, understanding interactions between HPVs and the immune system may shed new light on the factors that lead to clearance or chronicity, with implications for human cancers of infectious origin (*16*), and the development of immunotherapies (*17*). Finally, non-persisting HPV infections represent a major reservoir of virus diversity, which could fuel an evolutionary response to vaccine-driven selective pressures (*18*). Unfortunately, although we have known for decades about the prevalence and duration of non-persisting HPV infections (*4, 5, 19*), we still know little about the immune response they might elicit and temporal variations in virus loads.

To address this matter, we implemented a longitudinal cohort study in Montpellier (France) during which 189 women aged from 18 to 25 years old were followed every two months until HPV infection clearance or for a maximum duration of 24 months (*20*). At each of the 974 on-site visits, biological samples were collected and participants filled in detailed socio-demographic, health, and behaviour questionnaires. The strength of the study stems from its temporal resolution and the quality of the biological data generated. The main characteristics of the cohorts are shown in Table S1 and in Ref. (*21*).

## Immune response to HPV infections

HPV infections are poorly immunogenic and only 40 to 60% of HPV-positive women seroconvert to an incident viral genotype (*22*). Therefore, we focused on the local cellular immune response and used flow cytometry (FCM) to analyse cervical smears. These biological samples are particularly fragile and challenging to study because keratinocytes are highly adhesive and autofluorescent cells. Building on an existing protocol (*23*) and on existing software packages (*24*), we devised a pipeline to identify clusters of immune cell populations with a non-supervised approach. We first used this pipeline to identify leukocytes (CD45^+^ cells). We then applied it to this cell population to automatically delineate 20 cellular clusters, which we could manually group into 11 distinct immune cell populations based on morphological and lineage markers (Fig. S2, S3, and Table S3). The most frequent cells were CD16^+^ granulocytes (59.9%). We could also notably identify three distinct populations of TCRγδ cells (clusters III, VII and IX), representing respectively 3.88%, 12.44%, and 0.73% of the CD45^+^ cells (Fig S2), as well as CD4 and CD8 T cells (respectively 1.15% and 0.78%). Other cell populations could not be assigned formally but display features of local antigen-presenting cells (cluster V) or NK cells (cluster II) (Fig. S2).

We stratified the samples as HPV negative or positive for a ‘focal’ HPV genotype, *i*.*e*. a genotype detected at least during two consecutive visits (Fig. S1 and Table S2). This emphasis was made to avoid a spurious focus on ‘singletons’, *i*.*e*. an HPV genotype detected at a single visit. These are sometimes referred to as ‘transient infections’ (*25*) and are a poor marker of actual infections (*26*). A UMAP clustering helped visualise the 11 immune cell populations and the composition shift in samples with a focal HPV, especially an increase of cluster VII (Fig. 1A). A differential abundance analysis (Fig. 1B) confirmed that focal HPV-positive samples exhibited a lower proportion of cluster X (CD4^+^ T-cells) than HPV-negative samples (Fold Change, FC: 0.62) and a higher abundance of cluster VII (CD45^low^ TCRγδ cells) and cluster VIII (FC: 1.71 and 1.41, Fig. 1B and C). Clusters IV, V, and VI were also rarer in focal HPV-positive samples (FC: 0.63, 0.71, and 0.67). These differences were statistically significant in a multivariate generalized linear mixed model including anti-HPV vaccination status as a fixed effect and a random effect accounting for the multiple sampling per participant (Fig. 1C). Differential expression analysis highlighted moderate changes for activation markers CD69 or CD161 fluorescence levels. We did find a reduced median signal intensity for CD69 on CD4 T cells, which suggests lower priming of CD4 T cells during focal HPV infections (cluster X, Fold change = 0.90, p-adj = 0.04) (Table. S4).

**Figure 1.**
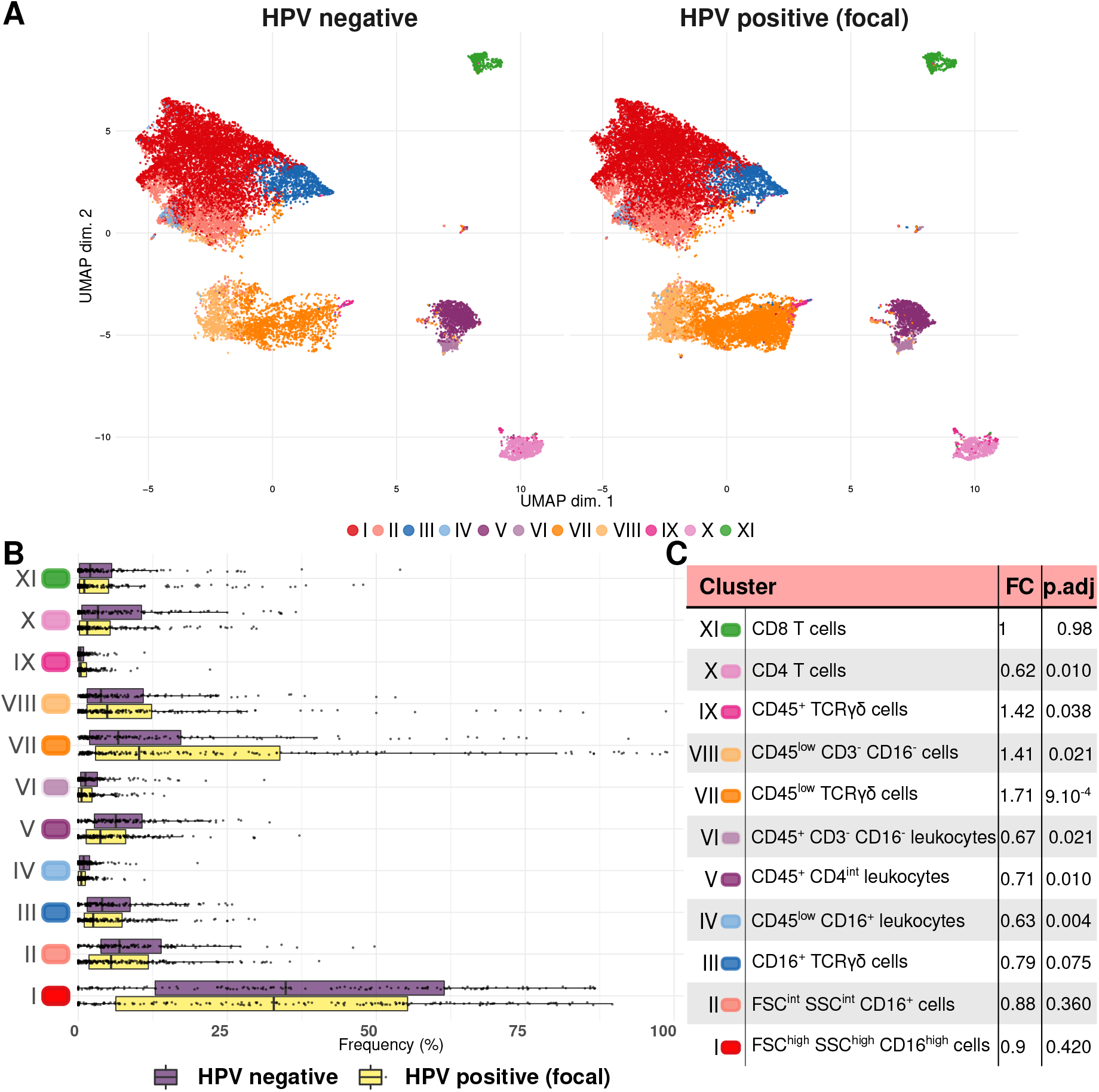
Unsupervised clustering of flow cytometry data from cervical smears. A) Highlighting 11 homogeneous populations using UMAP clustering on data from samples without HPV or positive for a ‘focal’ HPV infection, B) Comparison of the clusters frequencies based on infection status, and C) Cluster annotation and fold change (FC) values shown in panels A and B. FC were calculated by an abundance analysis with diffcyt-DA-edgeR adjusted with a Benjamini-Hochberg test.

Building on previous results showing an association with HPV infections (*27*), we quantified the concentrations of five cytokines in cervical secretions using Meso Scale Discovery technology and normalised the values over total protein concentration. Associations with HPV status (Fig. S4) were consistent with earlier cross-sectional studies (*27, 28*). To improve the characterization of immune cells, we explored correlations between the proportion of each immune cell population in a sample and the concentration of each cytokine in the same sample (Fig. 2A). Our linear models identified positive correlations between the concentration of IFNγ and the frequency of CD8 T cells (cluster XI, *β* = 0.14), although with a p-value of 0.062, and to a lower extent with CD45^+^ TCRγδ cells (cluster IX, *β* = 0.05, *p* = 0.03). We also found a negative correlation between IL-17A and cluster VIII (*β* = −0.28, *p* = 0.019).

**Figure 2.**
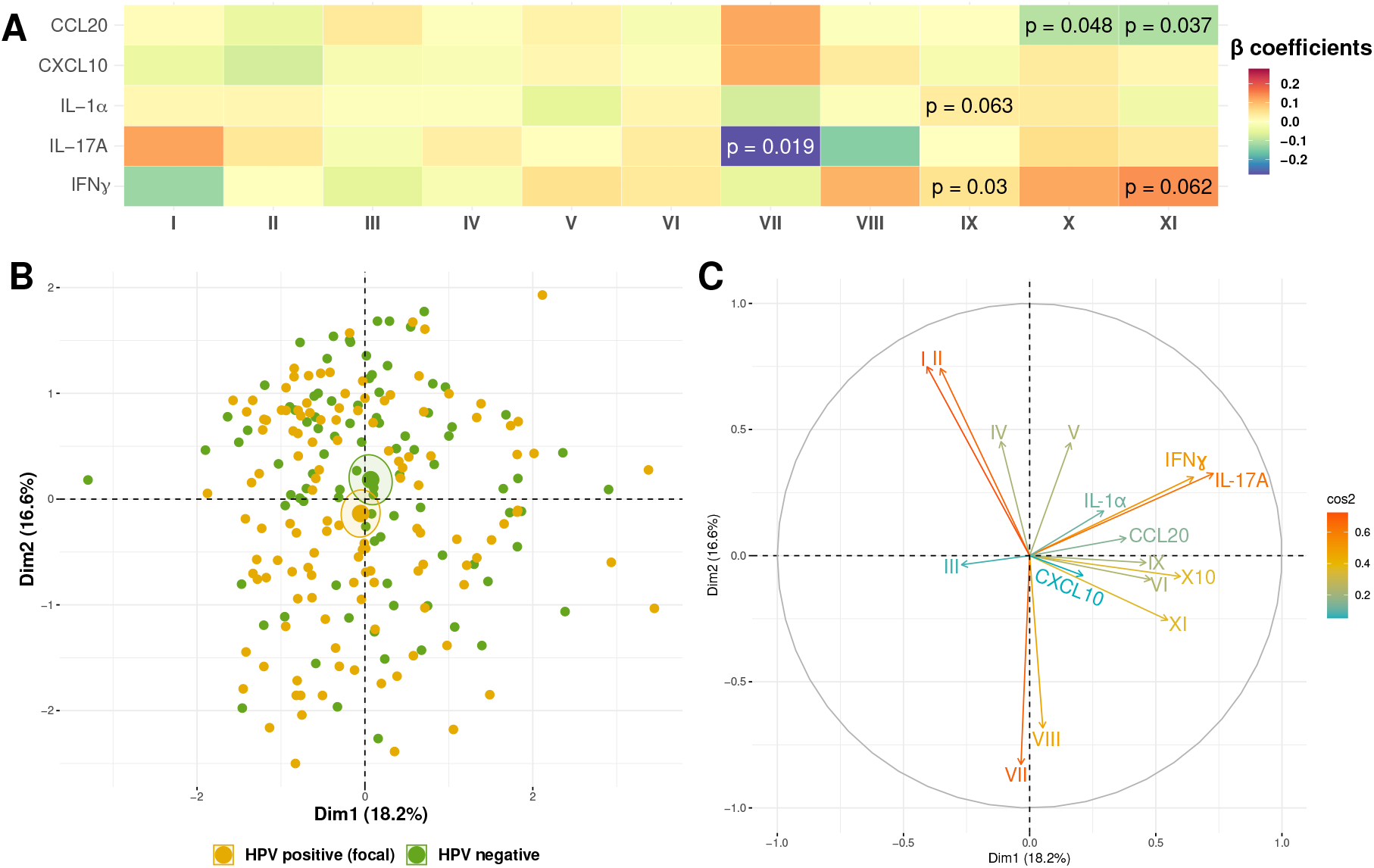
Local immune response in HPV-infected and uninfected women. A) Correlation matrix between the local density of five cytokines and the proportion of the 11 cell clusters from Figure 1. B) Sample clustering based on focal HPV infection status. C) Axes of a multiple factor analysis based on FCM and cervical cytokines and chemokines. In A, colors indicate the coefficient of pairwise regressions (only p-values lower than 10% are shown).

Combining all cervical immune variables (cytokines and immune cell populations) in a multiple factor analysis showed a trend for differential clustering of samples originating from HPV focal infections and from uninfected women (Fig. 2B). The axes indicate that HPV focal infections were negatively correlated with IFN-γ and IL-17a concentrations and frequency of CD16^+^ granulocytes (clusters I and II) (Fig. 2C). Conversely, the main vectors associated with HPV focal infection were CD45^low^ TCRγδ cells (cluster VII) and cluster VIII (Fig. 2C).

### Viral load kinetics

For all 126 participants with at least one HPV genotype detected, we estimated the virus load in all the cervical smears collected at each on-site visit for 13 HPV genotypes using a sensitive and specific quantitative Polymerase Chain Reaction (qPCR) protocol (*29*). The number of viral copies was normalised by that of a cellular gene (albumin). We could monitor 160 infections, including 28 complete infections (18%), 99 left-censored where participants were enrolled as positive (62%), and 69 right-censored where clearance was not observed (43%) (Fig. 3). Given the longitudinal nature of our data, we turned to the field of viral kinetics (*30*) and developed Bayesian non-linear mixed-effects models (*31, 32*). We described virus dynamics with five parameters capturing an increasing slope, a plateau, and a declining slope (Fig 4A and Methods). This shape was motivated by the important frequency of left- and right-censored follow-ups and the fast rate of reaching the plateau, which a simpler model with two slopes could not capture. Moreover, a similar plateau was observed to emerge in a previous mathematical model of HPV dynamics in squamous epithelia (*33*).

**Figure 3.**
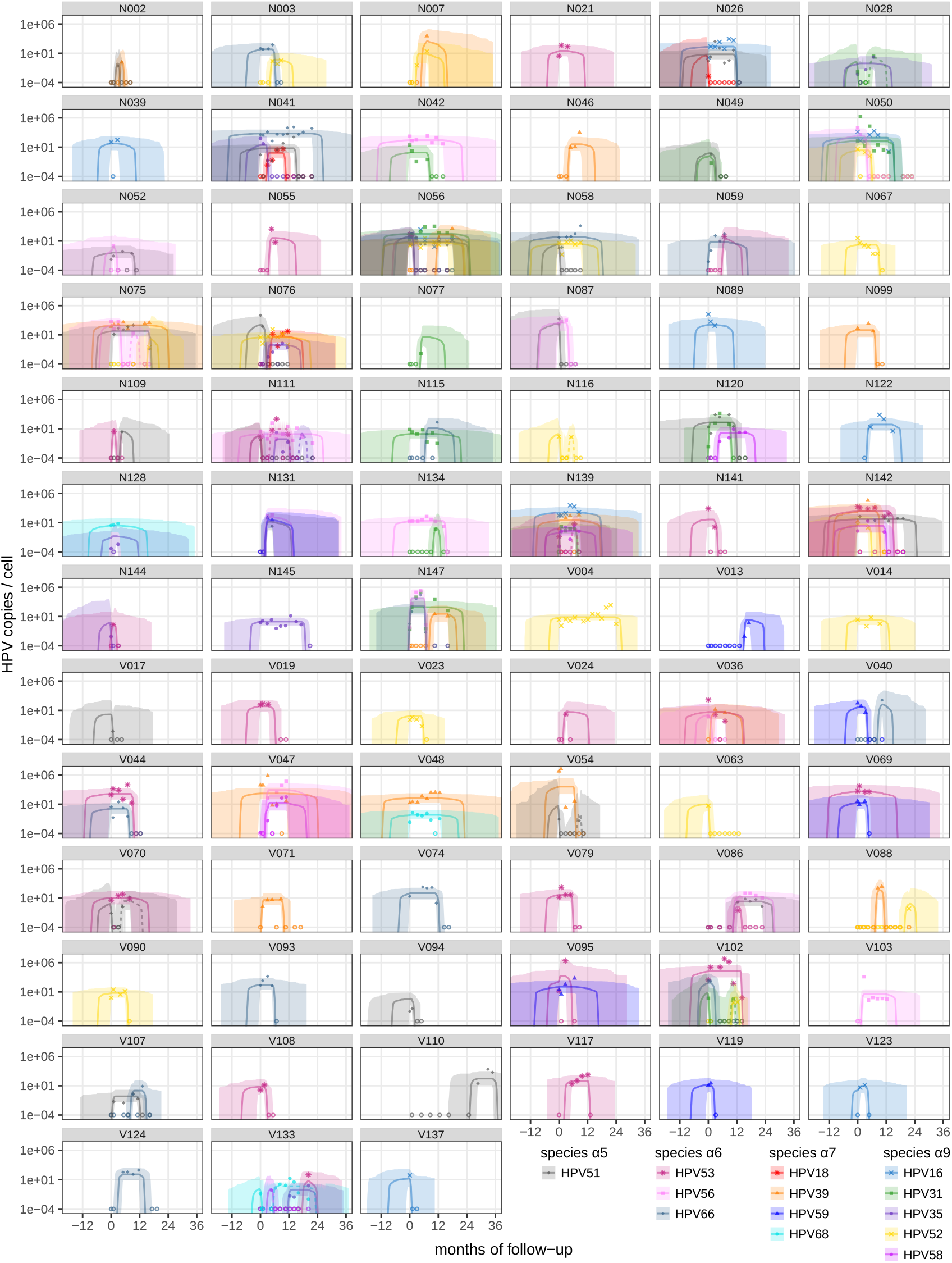
Virus load kinetics for 160 HPV genital infections in 75 women. Each panel corresponds to one participant and shows the number of HPV genome copies per number of human genome copies resulting from a three-slopes hierarchical Bayesian model. The lines show the posterior median trajectory and shaded area the 95% credibility interval. Open circles indicate values below the limit of detection. The letter before the anonymity number (above each panel) indicates whether the participant was vaccinated (V) or not (N).

**Figure 4.**
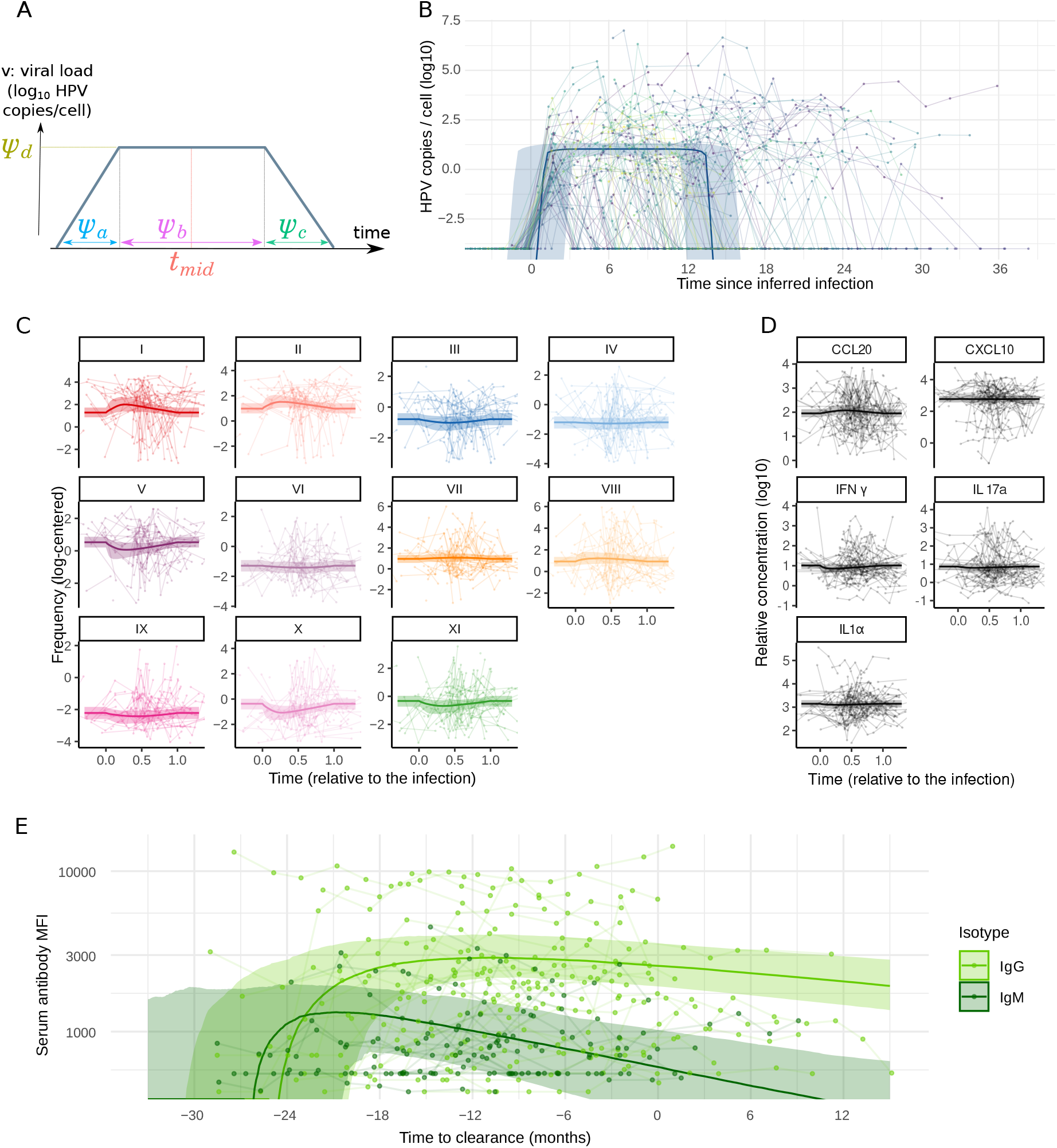
Population predictions of virus and immune kinetics in HPV infections. A) Representation of the parameters governing the descriptive models. The viral kinetics model is governed by five parameters: plateau virus load *ψ*_*d*_, time to the midpoint of the infection *t*_mid_, growth phase duration *ψ*_*a*_, plateau duration *ψ*_*b*_, and clearance phase duration *ψ*_*c*_. B) Population prediction of virus load dynamics. As in all the panels, the black thick line shows the median trajectory, the shaded area the 95% highest posterior distribution, each dot represents an observation, and thin lines show observed trajectories of individual infections. C) Population prediction for immune cell dynamics in HPV infections. Cell proportions are transformed with a centered log-ratio. D) Population prediction for cytokines dynamics in HPV infections. In panels C and D, time is normalized between 0 (infection) and 1 (clearance). E) Prediction of IgG (light green) and IgM (dark green) kinetics before and during HPV infections. MFI stands for mean fluorescence intensity.

The resulting model parameter estimates captured the per-participant variability in normalised virus load values and exhibited wider uncertainty in censored follow-ups (Fig. 3). The population average of the mixed effect model (Fig. 4B) indicated that infections lasted 14 months (95% Credibility Interval, CrI, 11 to 18), with a 12 months (95% CrI: 10 to 15) plateau at 11 virus genomic copies per cell (95% CrI: 5 to 21). According to the model, 90% of the infections clear within three years, which is in line with earlier studies (*7*).

### Immune kinetics

We then used the same method but with a different model to analyse the flow cytometry (FCM) time series. We transformed the cell population proportions using centered log-ratios (*34*) and assumed a two-slope model with matching dates of the beginning and end of the infection to the virus load fit. At the population level, some clusters of cells exhibited similar kinetics during the course of infection (Fig. 4C). The first group consisted of clusters I and II, which we associated with the innate immune response, and increased in relative proportion after the onset of infection. Another group (clusters VII and VIII) exhibited a smaller and later peak. Finally, a third group (clusters VI, IX, X, and XI) was associated with the adaptive immune response and decreased in frequency over the course of the infection. Moreover, the frequency of the remaining clusters (III, IV, and V), which were associated with the innate immune response, tended to decline during the infection. These cells clusters groups also appear when analysing inter-individual variation through the random effects correlation matrixes inferred by the model (Fig. S7).

We assumed the same two-slope model for the cytokines concentration dynamics in the cervix. At the population level, CCL20 increased over the course of the infection whereas IFNγdecreased. The other cytokines remained steady, although with an important variability between participants (Fig. 4D). Analysing inter-individual variation patterns indicated that the concentrations of CCL20 and CXCL10 were closely correlated, and negatively correlated to those of IL-17A and IFNγ (Fig. S8).

Finally, circulating IgG and IgM antibodies specifically targeting 10 HPV genotypes were quantified using a multiplex assay (*21, 35*). We assumed a classical Bateman function for the dynamics (*36*) and only considered IgG and IgM specific to the infecting HPV genotype. This limited the analysis to 55 infections in 37 individuals but still allowed us to identify a marked difference between IgG and IgM with a steeper decline of the latter (Fig. 4E).

### Linking viral dynamics and immunity

We then compared the infection duration and the magnitude of the virus load with the immune response. For multiply infected individuals, we focused on the longest infection, which also generally had the highest plateau viral load (Fig. S6). Because of the compositional (and therefore highly correlated) nature of the data, we used partial least square (PLS) regressions for the analysis.

The regression explained a third of the variance in infection duration 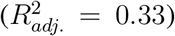, which was found to be significantly correlated with the mean concentration of IFNγ and the mean frequency of FCM clusters VIII and IX, but negatively associated with CXCL10 (Fig. 5A). Furthermore, HPV species α5 had a significantly shorter infection duration than the reference species α9 (which includes HPV16). The strong association between the multiply-infected status and the infection duration was expected since we kept the longest infection per participant in the model. Finally, for a smaller subset of participants infected by a genotype we could identify both with serology and qPCR, being seropositive in IgGs beforehand led to shorter infections (9.9 months, 95% CI: 7.7-12.8) than in seronegative individuals, even if they seroconverted (17.0 months, 95% CI: 12.0-24.1, Fig. 5B). Out of the 12 infection events where we observed the end of the infection and the individuals were initially seronegative, four never seroconverted (33%).

**Figure 5.**
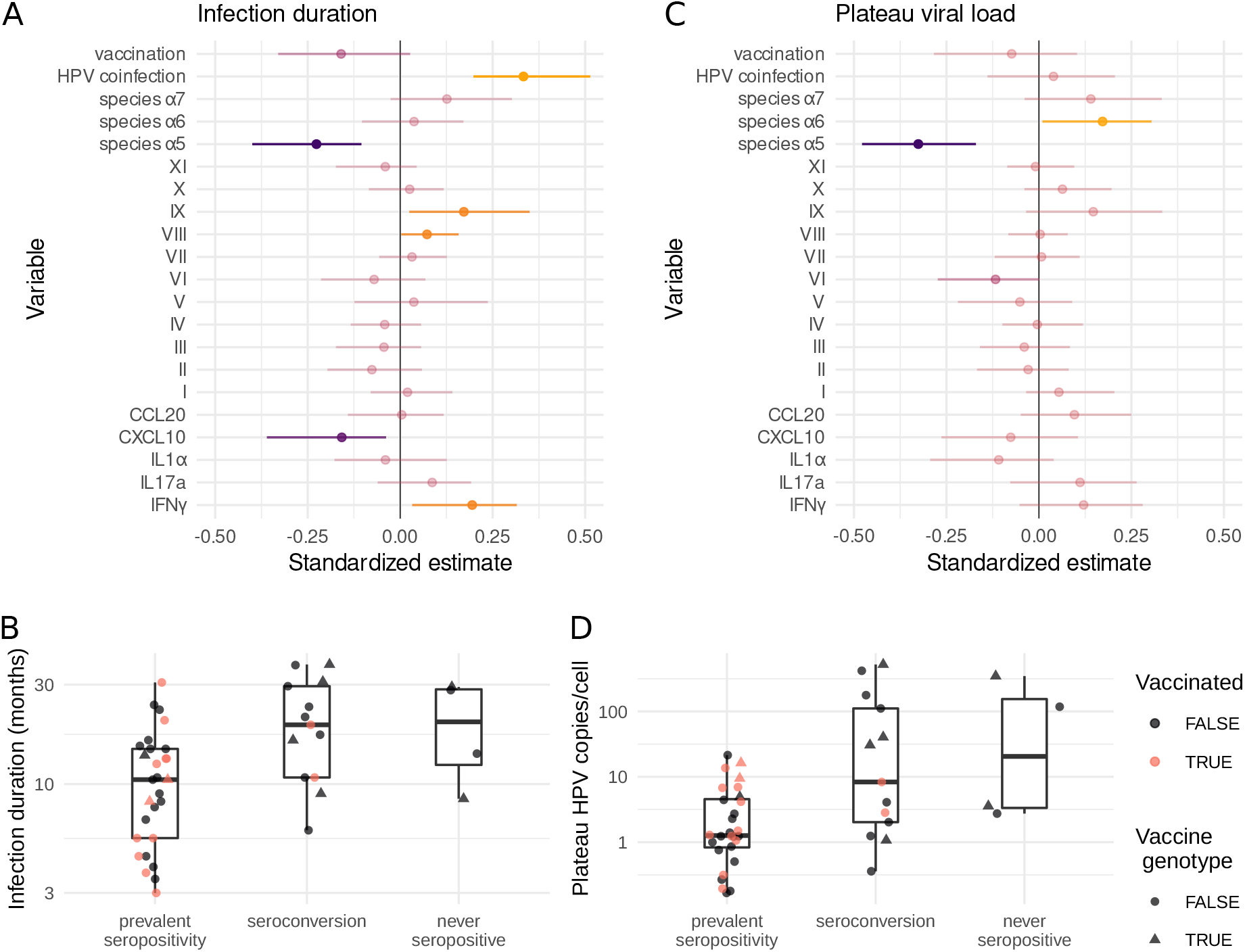
Factors associated with HPV infection duration and plateau viral load. A) Outcome of partial least squares (PLS) regressions between the infection duration and the immune response summary statistics 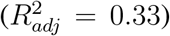, and B) between the viral load during the plateau and the immune response summary statistics 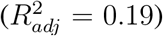. Variables with a 95% CI different from 0 are shown in brighter colors (yellow for a positive correlation and purple for a negative one). The HPV reference species is α9, which includes HPV16. C) Association between the inferred timing of seropositivation on infection duration and D) plateau virus load. This timing has a significant effect on the infection duration (*F* [2, 37] = 4.3, *p* = 0.023) and on the plateau viral load (*F* [2, 41] = 7.6, *p* = 1.5 10^−3^). Colors indicate vaccination status and shapes the genotype status (*i*.*e*. HPV16 or HPV18).

Regarding the magnitude of the plateau virus load, the proportion of the variance explained by the PLS regression was lower than for the infection duration 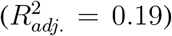 (Fig. 5C) but the HPV species had a stronger impact. Indeed, compared to the reference species α9, α6 had a higher viral load, whereas α5 had a lower one. The mean frequency of cluster VI was also negatively correlated with the plateau viral load (although only with *p <* 0.1). This is consistent with this cluster most likely corresponding to B lymphocytes (our FCM analysis being supported by the observation that among women with matching IgG data, its frequency was associated with the mean IgG titers in the serum, Fig. S9). Finally, women who were seropositive before the infection had a plateau viral load on average 7.6 times smaller (95% CI: 2.4-25) than those who seroconverted during the infection (Fig. 5D).

## Discussion

While most HPV infections in young adults last less than two years (*4, 6, 7, 15*), HPV clearance processes and mechanisms remain elusive. Thanks to the combination of dense monitoring, high-quality data, and statistical models, we offer original insights into HPV-immunity clearance dynamics.

HPV infections exhibit three phases: a short growth phase, a long plateau phase, and, for some of them, a short clearance phase. This is consistent with an existing mathematical model (*33*) and prior knowledge on infection duration. Our work is limited to studying the path to chronic infections because participants were not followed after 24 months of infection by the same HPV (only 4 of the 126 infected women did not clear the infection in 18 months). Furthermore, although dense, our follow-up has a 4 months uncertainty window around ‘singletons’, i.e. HPV detection at a unique visit. Studies with denser follow-ups are required to better understand whether these events correspond to short productive infections.

Given the little prior knowledge, we studied both innate and adaptive immune responses. Focusing on lymphoid immunity and harnessing the power of unsupervised clustering enabled us to highlight the importance of TCRγδ cells on HPV dynamics, with at least three distinct subpopulations. The dominance of TCRγδ cells in mucosal immunity could explain their relative abundance in our samples. This is also consistent with recent studies on cervical smears that did not include these cells in their panels but found a relatively higher abundance of T cells (*37, 38*).

We also found a lower proportion of CD4 T cells (cluster X) in individuals with a focal HPV infection compared to uninfected individuals. Furthermore, a population yet to be characterised in details, cluster VIII, was more frequent in focal HPV infections and associated with longer infections. Based on its expression pattern, we hypothesise that it could contain ILCs or progenitors, but not ILC3 given the lack of correlation between this cluster and IL-17A (Fig. 2) (*39*).

Among the infected participants for whom we could monitor viral kinetics by qPCR, Bayesian hierarchical modelling identified a positive correlation between the infection duration and only one of the three TCRγδ populations (cluster IX with CD45^+^ TCRγδ cells). This highlights the diversity of these populations and points to their different roles at specific times of the infection course (*40*).

Infection duration was also positively correlated with IFNγ mean concentration in the cervical area, which is consistent with earlier results. Unexpectedly, longer infections were associated with decreased concentrations of CXCL10, although the release of this cytokine is usually stimulated by IFNγ (*41*). One interpretation is that averaging the overall response during the infection may buffer time-dependent patterns of the antiviral response. Another hypothesis could be that only the HPV infections that manage to evade the innate immune response can establish a more persistent infection, thereby triggering the adaptive immune response. In this case, CXCL10 would only be elevated during the early immune response. This is consistent with results from a comparable cohort indicating that persistence is associated with high levels of proinflammatory, Type-1, and regulatory cytokines (*25*).

We did not observe any ‘breakthrough’ focal infection in the study and, as a consequence, vaccine status was not associated with infection duration or plateau viral load. We did, however, observe a trend towards shorter infections among vaccinated individuals, which could be attributed to a mild cross-protective mechanism. This is also consistent with our observation that participants who were seropositive for an HPV genotype before being infected by this same genotype exhibited shorter infections with lower virus loads. This shows that natural immunity may provide protection against infection but is not sufficient to prevent it. Note that this importance of the serological status, which was missing for some HPV types, imposes nuancing the role of the other factors on the duration of the infection and the plateau virus load.

Our immunological analyses lay the ground for future work. Future extensions could be to investigate markers more related to myeloid immune cells. On the statistical side, adding a temporal dimension via the kinetics analyses allowed us to extract more information from the data by harnessing the longitudinal information inherent to the cohort. Future studies could build on existing within-host dynamics models to estimate more biologically-relevant parameters, such as the ‘burst size’ of infected cells or the killing rate by the immune response (*33*).

Both the vaginal microbiota (*42*) and the immune response vary over the course of the menstrual cycle (*43*) with, for instance, fewer immunoglobulins during the luteal phase and, conversely, more IL-1α. Furthermore, the vaginal community state type has been associated with the risk of HPV detection (*26*) but investigating how menstrual cycles interact with the course of HPV infection dynamics would require denser follow-ups.

Cohort participants were followed for a mean duration of 290 days. Long-term follow-up of these women could yield valuable insights, especially thanks to new development in HPV full genome sequencing (*44*). For instance, it could help to estimate the prevalence of latent infections (*45*) but also further monitor the dynamics of chronic infections (*46*). Finally, it would provide critical data on HPV within-host evolution, which could be associated with cancer development since HPV16 found in cervical cancers has a particular genomic signature with less variation in the E7 gene (*47*).

Beyond the case of HPVs, this system represents an opportunity to better understand the sometimes tenuous frontier between non-persisting and chronic viral infections (*48*).

## Supporting information

Supplementary Materials Tessandier & Elie

## Data Availability

The raw data and R scripts used will be deposited on the Zenodo server upon publication.

## Acknowledgments

This project has received funding from the European Research Council (ERC) under the European Union’s Horizon 2020 research and innovation programme (grant agreement No 648963, to SA).

The authors acknowledge further support from the Centre National de la Recherche Scientifique, the Institut de Recherche pour le Développement, the Fédération Hospitalière Universitaire InCH of Montpellier, the Fondation pour la Recherche Medicale (to TK), the Ligue contre le Cancer (to TB), and the Agence Nationale de la Recherche contre le Sida (ANRS-MIE, to NT and OS).

The authors acknowledge the ISO 9001 certified IRD i-Trop HPC (member of the South Green Platform) at IRD Montpellier for providing HPC resources that have contributed to the research results reported within this article (bioinfo.ird.fr and www.southgreen.fr).

The raw data and R scripts used will be deposited on the Zenodo server upon publication.

## Supplementary materials

Materials and Methods

Supplementary Results

Figures S1 to S9

Tables S1 to S9

References (S1-S17)

Ethics, competing interests, and authors’ contributions

