## Supplementary Materials Tessandier & Elie for "Viral and immune dynamics of HPV genital infections in young women"

### Supplemenatry Materials

**Supplementary References** **57**

### S1 Materials & Methods

#### S1.1 The PAPCLEAR clinical study

The PAPCLEAR clinical study protocol is summarised below and additional details can be found in Ref. (S*1*). This monocentric cohort study followed $N = 149$ women longitudinally between 2016 and 2020. Its inclusion criteria were to be between 18 and 25 years old, to live in the area of Montpellier (France), to be in good health (no chronic disease), not to have a history of HPV infection (e.g. genital warts or high-grade cervical lesion), and to report at least one new sexual partner over the last 12 months. These criteria were chosen to maximise the probability of HPV infection acquisition over the follow-up. Enrollment was independent of HPV infection and HPV vaccination status.

Participants were enrolled by putting up posters and handing out leaflets at the main STI detection centre (CeGIDD) within the University Hospital of Montpellier (CHU) and in the Universities of the city. To increase enrolment, posters were also hung at bus stops near the CHU.

The inclusion visit (V1) was performed by a gynaecologist or a midwife at the CeGIDD outside operating hours. After an interview, several samples were collected by the clinician (vaginal swabs, ophthalmic sponges, and cervical smears) and are further described below. A nurse then collected blood samples. Finally, participants filled in a detailed questionnaire.

A results visit (V2) was scheduled 4 weeks later to inform the participants of the result of a screening test for cervical lesions performed via liquid cytology (Thinprep medium from Hologic). Depending on their HPV status at the inclusion visit (see below), participants were oriented to the HPV-negative or to the HPV-positive arm of the study.

Participants in the HPV-negative arm, for whom no alphapapillomavirus was detected at V1, came back for on-site visits every 4 months. If an HPV was detected at one of the following visits, they switched to the HPV-positive arm. HPV-negative participants who were not infected by month 32 of the study were not followed anymore.

Participants in the HPV-positive arm had on-site visits every two months until clearance or chronification of the infection. Chronicity was defined as 24 months of infection by the same HPV genotype. Clearance of a specific infection was defined as two successive visits without any detection of the focal genotype.

To improve the statistical power of the cohort, this longitudinal study was complemented by a cross-sectional study, which was originally designed to enroll the same number of women, i.e. 150. Participants in the cross-sectional study only performed the inclusion visit (V1) and the results visit (V2).

The inclusions began in November 2016 and, due to the COVID-19 pandemic and the associated strain on hospital services, the study had to be prematurely ended in September 2020. This led to the loss of follow-up of two participants and 110 non-inclusion in the cross-sectional cohort. Overall, we followed $N = 189$ women, including 149 (78.8%) in the longitudinal cohort, with a total of 974 on-site visits with gynaecological consults.

#### S1.2 Clinical samples description and processing

##### S1.2.1 Cervical smears

A cervical smear was collected at each of the on-site visits. The gynaecologist or midwife performed 2.5 turns before putting the brush into 20mL Thinprep™ PreserCyt™ medium from Hologic (at V1) or 5mL of fresh PBS medium (at the other visits).

Smears in PreserCyt™ medium were sent to the anatomical pathology department of the University Hospital of Montpellier for cytological analysis by a trained pathologist. Before that, 2mL were sampled for HPV detection. Leftovers from the cytological analysis were also stored.

Smears in PBS were processed following a general protocol described in Ref. (S2). The whole protocol was performed at 4°C. Within 2h of collection, the smears were vortexed during 45s with the cytobrush that was then removed carefully to save as many cells as possible and 5 mL of RPMI was added. The solution was then filtered in a new tube with a cell strainer (Fisherbrand™ Sterile Cell Strainers, 100µm). 250µL of the resulting solution were aliquoted for back up. The remaining solution was centrifuged for 10min. at 514g and the supernatant was stored at -80°C and the cell pellet was washed with 5mL PBS at 514g for 10min. The supernatant was discarded, and cells were resuspended in 200µL. 20µL of the resulting solution was aliquoted for HPV detection while the rest was processed for flow cytometry staining.

##### S1.2.2 Ophtalmic sponges

As described in Ref. (S*3*), during the on-site visits, cervicovaginal secretions were collected by the gynaecologist or midwife using WeckCel sponges (Beaver-Visitec International) placed directly into the cervical os for 1 minute. Sponges were then transferred into a Salivette® (Sarstedt) device and centrifugated at 1500rpm for 5 min at 4°C after the addition of 175µL of phosphate-buffered saline (PBS). Supernatants were separated into 50µL aliquots and stored at -80°C.

##### S1.2.3 Blood samples

During onsite visits, 20mL of circulating blood was collected by a trained nurse for antibody titration and STI detection.

#### S1.3 HPV detection and genotyping

For HPV detection and typing, we started from the sample originating from the cervical smear and resuspended in 200µL of PBS as described above. From this, we extracted DNA using the QIAamp® DNA mini kit (QIAGEN Inc.) following standard protocol for body fluids (spin control).

We first tested for the presence of alphapapillomaviruses using the generic DEIA test (S*4*) from

DDL Diagnostic Laboratory (Rijswijk, the Netherlands). We then used the $LiPA_{25}$ typing (S5), also from DDL Diagnostic Laboratory, on DEIA-positive samples. Both assays target the same amplicon of the L1 viral gene.

Samples that were DEIA-positive and $LiPA_{25}$ negative were amplified using the PGMY PCR (S6) and sequenced using Sanger sequencing. Samples for which the sequencing did not yield a clear sequence, most likely because of coinfections, were labelled as 'non-typable'.

#### S1.4 Genotype specific HPV qPCR

We quantified the number of genomic copies by quantitative Polymerase Chain Reaction (qPCR) for 12 HPV genotypes (HPV16, 31, 35, 39, 51, 52, 53, 56, 58, 59, 66, 68) using the protocols and primers from Ref. (S7). We also quantified the number of copies of HPV18 and of a human reference gene (albumin) using a protocol shared by Pr Pretet's team in Besançon, France. The HPV18 forward primer was 5'-ACACCACAATACCATGGCG-3', the HPV18 reverse primer was 5'- TTCAGTTCCGTGCACAGATC -3', and the HPV18 probe was 5'-FAM-CAACACG\GCGACCCTACAAGCTAC-HBQ1-3'. For the albumin primers, the details can be found in Ref. (S8).

All qPCRs were performed using SensiFAST™ Probe No-ROX Kit (MERIDIAN Bioscience) on LightCycler® 96 and LightCycler® 480 (Roche Diagnostics) for 384 wells plates. Cycles used for all genotypes and albumin were 95°C for 5min and 40 two-step cycles with 95°C for 10s and 60°C for 30s. Each sample was run in triplicate with 2µL of samples in a final volume of 10µL. To create HPV standards, we used plasmids graciously provided by the international HPV reference center from the Karolinska Instituet (https://www.hpvcenter.se). Each plasmid contained a full HPV genome for one of the 13 target genotypes. For the albumin standard, we used genomic female Human DNA (Promega). For each participant, we ran on the sample plate the qPCR for all samples, all genotypes previously identified by $LiPa_{25}$, and the albumin gene.

Virus load measures were taken in triplicates and we analysed the mean cycle threshold (Ct) value. HPV genotype-specific Ct values were normalised by that of the cellular gene (the albumin) known to be present at 2 copies per diploid genome.

#### S1.5 Cytokines quantification

We used the MSD U-plex Biomarker Group 1 (human) from Meso Scale Discovery (MSD, Rockville, MD, USA) to measure the concentration of five analytes identified in a previous study as being associated with HPV infection (*S3*): IFN-$\gamma$, IL-10, CCL20, IL-17A, and CXCL10. According to the manufacturer's instructions, we used 25 µL of specimen, which was obtained from the ophtalmic sponges as detailed above. Analyses were performed on a MESO QuickPlex SQ 120 reader.

To normalize cytokine quantitation, we measured total protein concentration in the same samples using the Invitrogen Qubit™ protein assay (Thermo Fisher Scientific) following the manufacturer's instructions.

To compute cytokine sample concentrations from the standard curves, we used a four-parameter logistic regression model implemented in R with the package minpack.lm.

#### S1.6 Antibody dosage

IgG and IgM antibodies targeting L1 proteins of high-risk HPV types 16, 18, 31, 33, 35, 45, 52 and 58, as well as low-risk HPV-types 6 and 11 were analysed with a multiplex serology assay using beads coated with recombinant glutathione s-transferase (GST) fusions proteins. The assay procedure has been previously described in detail (*S9*). The samples were tested at a final serum dilution of 1:100 using an IgG and an IgM goat anti-Human secondary antibody. Seropositivity was defined based on standard definitions (*S10*, *S11*).

#### S1.7 Flow cytometry

For the flow cytometry (FCM) analyses, within three hours upon sampling, cellular suspensions were labelled with Live Dead red diluted 1/2000 (Thermofisher™) for 20 min in the dark, on ice. Cells were then washed with 2mL of PBS (1,500RPM, 5min) and resuspended in 100µL of PBS, before being transferred to a new tube containing dried antibodies (DURAClone, Beckman Coulter™). This custom antibody panel included anti-CD45 KRO (clone J33, IM2473U), anti-CD16 FITC (clone 3G8, B49215), anti-CD3 APC A750 (clone UCHT1, A94680), anti-CD4 APC A700 (clone 13B8.2, B10824), anti-CD8 PB (clone B9.11, A82791), anti-TCRγδPC5.5 (clone IMMU510, A99021), anti-CD69 PE (clone TP1.55.3, IM1943U), and anti-CD161 PC7 (clone 191B8, B30631).

Cells were incubated in presence of antibodies for 15min in the dark at room temperature, then washed with 2mL of PBS (1,500RPM, 5min). Finally, cells were re-suspended in 250 µL of PBS 1% PFA and stored at 4°C until acquisition using a flow cytometer (Navios, Beckman Coulter™).

#### S1.8 Unsupervised FCM analysis

For unsupervised analyses, raw LMD files were converted to FCS 3.0 using a custom R script. Samples with visible blood pellets were excluded from the analysis.

Raw files were cleaned using the PeacoQC R package, and then filtered for viable cells (LiveDead $R^-$). Using the CATALYST pipeline and FlowSoM clustering, we first restricted the dataset to $CD45^+$ cells.

We then clustered the cell populations using FlowSoM with 100 clusters and 20 metaclusters. The following parameters were used to perform the clustering: SSC-A, FSC-A, CD45, CD3, CD4, CD8, TCRγδ and CD16. CD69 and CD161 were only used for differential expression analysis. The 20 metaclusters thus generated were manually merged into 11 cell populations (see Figure S3 and Table S3).

A UMAP restricted to 150 cells per sample and on the same parameters used with FlowSoM clustering was performed to obtain a visual representation of the 11 cell population clusters.

We tested for differential abundances of the 11 cell populations between HPV-negative and focal HPV-positive samples using the diffcyt R package with the diffcyt-DA-edgeR method and including a random effect at the participant level. Differential expressions were calculated in a similar way using the diffcyt-DS-limma method.

#### S1.9 Basic statistical analyses

All statistical analyses were run with R.

In Table 1, we used Kruskal-Wallis rank sum test (`kruskal.test` function from the TableOne R package).

In Figure 2A, we calculated correlation matrices with the `lmer` coefficient (using the lme4 R package), assuming a random effect for the participant.

In Figure S4, we used the `glmer` function from the lme4 R package, with HPV positive (focal) as a response, the concentration of the five cytokines as covariates, and a random effect for the participant.

In Figure 2B and C, we performed a Mixed Factor Analysis (MFA) using the FactoMiner and FactoExtra R packages and including FCM frequencies (centered log-ratios) and the five cytokines and chemokines concentrations as variables.

#### S1.10 Bayesian hierarchical modeling

All the kinetics were modelled using RStan v. 2.21.5 and the script used is provided as Supplementary Material (the raw data will also be provided upon publication). In the following, we describe the statistical modelling approach performed to infer the viral dynamics and the associated immune dynamics.

The credible intervals were computed using the $2.5^{\text{th}}$ and the $97.5^{\text{th}}$ percentiles of the posterior distributions, *i.e.* with equally tailed intervals.

##### S1.10.1 Virus load dynamics

This analysis included all the participants who had at least three on-site visits and at least one positive virus load with the genotype-specific qPCR (see Fig. S1).

For some participants, for a given genotype, there were HPV-negative visits in-between HPV-positive visits. In the analysis, we assumed that HPV-positive visits separated by 2 or more consecutive negative visits (i.e. more than 4 months) corresponded to two different infection events. Overall, this generated 10 additional infections. We also conducted a sensitivity analysis to set this threshold to at least 3 negative visits.

As a quality check, samples with less than 100 copies of albumin were discarded from the analysis. Furthermore, following previous studies (S*12*), we set the limit of detection of HPV to $10^{-4}$ viral copies per cell.

Because of the shape of the raw data and of previous mechanistic mathematical modelling of HPV genital infections (S*13*), we assumed that virus load dynamics followed a pattern with an exponential growth phase, a plateau, and a rapid clearance phase. These dynamics were captured using five parameters (Fig 4A) describing the $\log_{10}$ virus load, i.e. the number of HPV copies per cell, over time $v(t)$ as follows:

$$v(t) = \begin{cases} v_0 & \text{if } t < t_{\text{mid}} - \psi_b/2 - \psi_a \\ v_0 + \frac{\psi_d - v_0}{\psi_a}(t - t_{\text{mid}} + \psi_b/2 + \psi_a) & \text{if } t < t_{\text{mid}} - \psi_b/2 \\ \psi_d & \text{if } t < t_{\text{mid}} + \psi_b/2 \\ \psi_d - \frac{\psi_d - v_0}{\psi_c}(t - t_{\text{mid}} - \psi_b/2) & \text{if } t < t_{\text{mid}} + \psi_b/2 + \psi_c \\ v_0 & \text{if } t > t_{\text{mid}} + \psi_b/2 + \psi_c \end{cases}$$

where, as illustrated in Figure 4 of the main text, the parameters indicate the plateau virus load $\psi_d$, the time to the midpoint of the infection $t_{\text{mid}}$, the growth phase duration $\psi_a$, the plateau duration $\psi_a$, and the clearance phase duration $\psi_a$. $v_0$ represents the minimal viral load and was chosen to enable satisfying convergence of the model.

Furthermore, in our analysis, we assumed two (nested) random effects: one for the participant ($i$) and one for each infection within this host ($j$). We did not include random effect parameters for the growth and the clearance phases of the dynamics because they were often unobserved due to the frequency of left-censored and right-censored follow-ups. Furthermore, even when observed, these phases were generally short compared to our sampling interval of two months.

Overall, we estimated the model parameters with the following assumptions:

$$\begin{aligned}
\psi_{a_{i,j}} &= \mu_a \\
\text{logit}\left(\frac{\psi_{b_{i,j}}}{\psi_{b_{\text{max}}}}\right) &= \text{logit}\left(\frac{\mu_b}{\psi_{b_{\text{max}}}}\right) + \eta_{b_i} + \rho_{b_j} \\
\psi_{c_{i,j}} &= \mu_c \\
\psi_{d_{i,j}} &= \mu_d + \eta_{d_i} + \rho_{d_j} \\
t_{\text{mid}_{i,j}} &= \mu_{\text{mid}} + \eta_{\text{mid}_i} + \rho_{\text{mid}_j}
\end{aligned}$$

where $\mu$s indicate the fixed effects,

$\begin{bmatrix}\eta_b \\ \eta_d\end{bmatrix} \sim N(0, \Omega_1)$, $\eta_{t_{\text{mid}}} \sim N(0, \omega_{1,t})$ are the between-host random effects, and

$\begin{bmatrix}\rho_b \\ \rho_d\end{bmatrix} \sim N(0, \Omega_2)$, $\rho_{t_{\text{mid}}} \sim N(0, \omega_{2,t})$ are the between-infections random effects.

$\Omega_1$ and $\Omega_2$ denote the variance-covariance matrix, with diagonal terms $\omega_1$ and $\omega_2$ respectively.

In our Bayesian analysis, we assumed the following priors for the parameters:

$$\mu_a \sim \mu_b \sim N(1, 0.5)$$
$$\mu_b \sim N(12, 4)$$
$$\mu_d \sim N(5, 1)$$
$$\mu_e \sim N(5, 4)$$
$$\omega_1 \sim \omega_2 \sim N((1, 0.5, 4), (0.5, 0, 3))$$
$$\sigma_{\text{vl}} \sim N(1, 0.25)$$
$$\Omega_1 \sim \Omega_2 \sim \text{LKJcorr}(2)$$

where LKJcorr indicates the Lewandowski, Kurowicka, and Joe correlation distribution (S*14*). We computed the likelihood assuming a mixture distribution of a normal additive error $\sigma_{vl}$ and a false-negative probability $\alpha$, which was fixed to 5% following earlier studies (S*15*). We ran the model using 4 chains with a maximal treedepth of 14. We used 15,000 iterations as a burn-in and sampled the next 3,000 iterations per chain to ensure having a minimal estimated sample size (ESS) of each parameter above 100.

###### S1.10.2 Immune dynamics: cytokines and FCM

In a similar way, but separately, for the $\log_{10}$ of the concentration of the cytokines and the log-ratio transformed cells frequencies, we assumed a two-slope dynamics through time, noted $c(t)$. We used the median posterior distribution of the virus dynamics to define the beginning and end of each infection, noted respectively $t_{\text{inf}}$ and $t_{\text{clear}}$, which also correspond to the beginning and the end of the slopes. Mathematically, we can write:

$$c(t) = \begin{cases} \psi_e & \text{if } t < t_{\text{inf}} \\ \psi_e + (t - t_{\text{inf}}) \frac{\psi_f}{(t_{\text{clear}} - t_{\text{inf}})\psi_g} & \text{if } t < t_{\text{inf}} + (t_{\text{clear}} - t_{\text{inf}})\psi_g \\ \psi_e + \psi_f - (t - t_{\text{inf}} - (t_{\text{clear}} - t_{\text{inf}})\psi_g) \frac{\psi_f}{(t_{\text{clear}} - t_{\text{inf}})(1-\psi_g)} & \text{if } t < t_{\text{clear}} \\ \psi_e & \text{if } t > t_{\text{clear}} \end{cases}$$

where $\psi_e$ was the basal value (before and after the infection), $\psi_f$ is the relative increase or decrease to this basal value, and $\psi_g$ is the relative timing of the peak (as a proportion of the total infection duration).

Overall, for each individual $i$ and each cytokine (or cell cluster) $j$, we estimated the following parameters:

$$\psi_{e_{i,j}} = \mu_{e_j} + \eta_{e_{i,j}}$$

$$\psi_{f_{i,j}} = \mu_{f_j} + \eta_{f_{i,j}}$$

$$\text{logit}\left(\frac{\psi_{g_{i,j}}}{\psi_{g_{\text{max}}}}\right) = \text{logit}\left(\frac{\mu_{g_j}}{\psi_{g_{\text{max}}}}\right) + \eta_{g_{i,j}}$$

where $\mu$s indicate the fixed effects and $\eta \sim N(0, \omega_3)$ the between-individuals random effect. We computed the likelihood assuming a normal distribution with an additive error $\sigma$.

For the FCM analysis, we assumed the following priors:

$$\mu_e \sim \mu_f \sim N(0, 1)$$

$$\mu_g \sim \text{Beta}(1.5, 1.5)$$

$$\omega_3 \sim N(0, 1.5)$$

$$\sigma_{\text{fcm}} \sim N(0, 1)$$

For the analysis of the cytokine, the priors were the following:

$$\begin{aligned}
\mu_e &\sim N(1.95, 1) \\
\mu_f &\sim N(0, 1) \\
\mu_g &\sim \text{Beta}(1.5, 1.5) \\
\omega_3 &\sim N(0, 1) \\
\sigma_{\text{cyt}} &\sim N(0, 1)
\end{aligned}$$

##### S1.10.3 Immune dynamics: antibodies

For the HPV-specific antibodies, given their expected dynamics, we assumed a classical Bateman function $a(t)$ with four parameters:

$$a(t) = \begin{cases} 0 & \text{if } t < \psi_m \\ \psi_l \frac{\psi_h}{\psi_h - \psi_k} (e^{-\psi_k (t-\psi_m)} e^{-\psi_h (t-\psi_m)}) & \text{else} \end{cases}$$

where $\psi_h$ is the antibody titer creation rate, $\psi_k$ the clearance rate, $\psi_l$ governed the peak titer, and $\psi_m$ represents the delay between the viral clearance and the start of the antibody response. We assumed that different genotypes infecting the same individual to be independent. Therefore, for each infection $i$, we estimated the following parameters:

$$\begin{aligned}
\log(1/\psi_{h_i}) &= \log(\mu_h) + \eta_{h_i} \\
\log(1/\psi_{k_i}) &= \log(\mu_k) + \eta_{k_i} \\
\log(\psi_{l_i}) &= \mu_l + \eta_{l_i} \\
t_{\text{clear}} - \log(\psi_{m_i}) &= \log(\mu_m) + \eta_{m_i}
\end{aligned}$$

Where $\mu$s indicate fixed effects and $\eta \sim N(0, \omega_4)$ the between-individuals random effect. We computed the likelihood assuming a normal distribution with an additive error $\sigma_1$ and a proportional term $\sigma_2$.

For the IgG analysis, the priors were the following:

$$\mu_h \sim N(3, 3)$$
$$\mu_k \sim N(36, 15)$$
$$\mu_l \sim N(8, 2)$$
$$\mu_m \sim N(18, 12)$$
$$\omega_4 \sim N(0, 4)$$
$$\sigma_{1,\text{IgG}} \sim N(0, 50)$$
$$\sigma_{2,\text{IgG}} \sim N(0, 1/3)$$

For the IgM analysis, the priors were the following:

$$\mu_h \sim N(0.5, 2)$$
$$\mu_k \sim N(18, 9)$$
$$\mu_l \sim N(7, 2)$$
$$\mu_m \sim N(12, 6)$$
$$\omega_4 \sim N(0, 2)$$
$$\sigma_{1,\text{IgM}} \sim N(0, 100)$$
$$\sigma_{2,\text{IgM}} \sim N(0, 1/4)$$

#### S1.11 Immune variables and viral kinetics regressions

For each immune response variable, we extracted the posterior median trajectory of each fit and computed the mean value during the whole infection period.

Due to the important correlation expected between the FCM variables due to their compositional nature, we used a partial least-squares (PLS) regressions to explore which variables were the most associated with the immune response and the plateau viral load.

These analyses were run using the R package plsRglm (S*16*) 95% credible intervals were inferred by performing 1,000 bootstraps and computing the adjusted bootstrap percentile (BCa) with the boot (S*17*) package.

The number of components was set following the AIC criterion with degrees of freedom computed with the dof package.

### S2 Supplementary Results

#### S2.1 Cohort profile stratified by HPV infection status

Table S1: **Cohort profile stratified by HPV infection status** Significant differences according to a t-test with a 5% significance threshold (unadjusted p-values) are shown in bold font. Table S2 shows the same variables but for HPV-focal infections only. Participants who were positive at least once in the follow-up were labelled as "HPV positive", whereas participants who were never positive during the follow-up were labelled as HPV negative.

| | HPV negative | HPV positive | p-value |
|---|---|---|---|
| number of participants ($n$) | 63 | 126 | |
| **Lifetime number of partners (mean (SD))** | 8.68 (8.41) | 12.06 (11.07) | 0.034 |
| **Vaccinated against HPV = Yes (%)** | 40 (63.5) | 57 (45.2) | 0.027 |
| Age at first visit (mean (SD)) | 21.43 (2.01) | 21.60 (2.02) | 0.576 |
| **Age at menarchy (mean (SD))** | 12.49 (1.23) | 12.87 (1.39) | 0.067 |
| First intercourse (age) (mean (SD)) | 16.62 (2.14) | 16.37 (1.85) | 0.401 |
| **Duration of follow up (days) (mean (SD))** | 198.30 (188.62) | 292.09 (213.02) | 0.003 |
| BMI (mean (SD)) | 21.99 (2.82) | 22.48 (3.62) | 0.352 |
| Antibiotics (last 2 weeks) = Yes (%) | 2 ( 3.2) | 10 ( 7.9) | 0.343 |
| Menses (last 2 weeks) = Yes (%) | 35 (55.6) | 64 (50.8) | 0.643 |
| Smoking (%) | | | 0.299 |
| No | 39 (61.9) | 82 (65.1) | |
| Occasionally | 12 (19.0) | 14 (11.1) | |
| Regularly | 12 (19.0) | 30 (23.8) | |
| Lubricant use (last 2 weeks) = Yes (%) | 12 (19.0) | 18 (14.3) | 0.526 |
| Intercourse with regular partner (last 2 weeks) = Yes (%) | 40 (63.5) | 70 (55.6) | 0.375 |
| Intercourse with occasional partner (last 2 weeks) = Yes (%) | 7 (11.1) | 18 (14.3) | 0.704 |
| Stress level (%) | | | 0.861 |
| 0 (Min) | 13 (20.6) | 20 (15.9) | |
| 1 | 25 (39.7) | 54 (42.9) | |
| 2 | 18 (28.6) | 39 (31.0) | |
| 3 (Max) | 7 (11.1) | 13 (10.3) | |

611 ## S2.2 Cohort profile stratified by focal HPV infection status

Table S2: **PAPCLEAR cohort profile stratified by HPV focal infection status.** Significant differences according to a t-test with a 5% threshold are shown in bold font (unadjusted p-values). Table S1 shows the same variables stratified by HPV status. Focal infections are defined as two consecutive visits for the same HPV. Participants with a transient infection are not included in this analysis (see Methods and Fig. S1).

| | No focal infection | Focal infection | p |
|---|---|---|---|
| number of participants (n) | 97 | 92 | |
| **Lifetime number of partners (mean (SD))** | 8.82 (8.21) | 13.16 (11.87) | 0.004 |
| **Vaccinated against HPV = Yes (%)** | 57 (58.8) | 40 ( 43.5) | 0.050 |
| Age at first visit (mean (SD)) | 21.63 (1.99) | 21.46 (2.05) | 0.559 |
| **Age at menarchy (mean (SD))** | 12.54 (1.34) | 12.97 (1.32) | 0.028 |
| **Duration of follow up (days) (mean (SD))** | 195.91 (180.04) | 329.27 (217.27) | <0.001 |
| BMI (mean (SD)) | 22.42 (3.28) | 22.21 (3.49) | 0.662 |
| **First intercourse (age) (mean (SD))** | 16.68 (2.04) | 16.21 (1.83) | 0.095 |
| Antibiotics (last 2 weeks) = Yes (n (%)) | 5 ( 5.2) | 7 ( 7.6) | 0.694 |
| Menses (last 2 weeks) = Yes (n ((%)) | 55 (56.7) | 44 ( 47.8) | 0.282 |
| Smoking (n ((%)) | | | 0.426 |
| No | 62 (63.9) | 59 ( 64.1) | |
| Occasionally | 16 (16.5) | 10 ( 10.9) | |
| Regularly | 19 (19.6) | 23 ( 25.0) | |
| Lubricant use (last 2 weeks) = Yes (n ((%)) | 17 (17.5) | 13 ( 14.1) | 0.660 |
| **Intercourse with regular partner (last 2 weeks) = Yes (n ((%))** | 63 (64.9) | 47 ( 51.1) | 0.074 |
| Intercourse with occasional partner (last 2 weeks) = Yes (n ((%)) | 11 (11.3) | 14 ( 15.2) | 0.568 |
| Stress level (last 2 weeks) (n ((%)) | | | 0.548 |
| 0 (Min) | 18 (18.6) | 15 ( 16.3) | |
| 1 | 42 (43.3) | 37 ( 40.2) | |
| 2 | 25 (25.8) | 32 ( 34.8) | |
| 3 (Max) | 12 (12.4) | 8 ( 8.7) | |

#### S2.3 Flow diagram of the sample selection

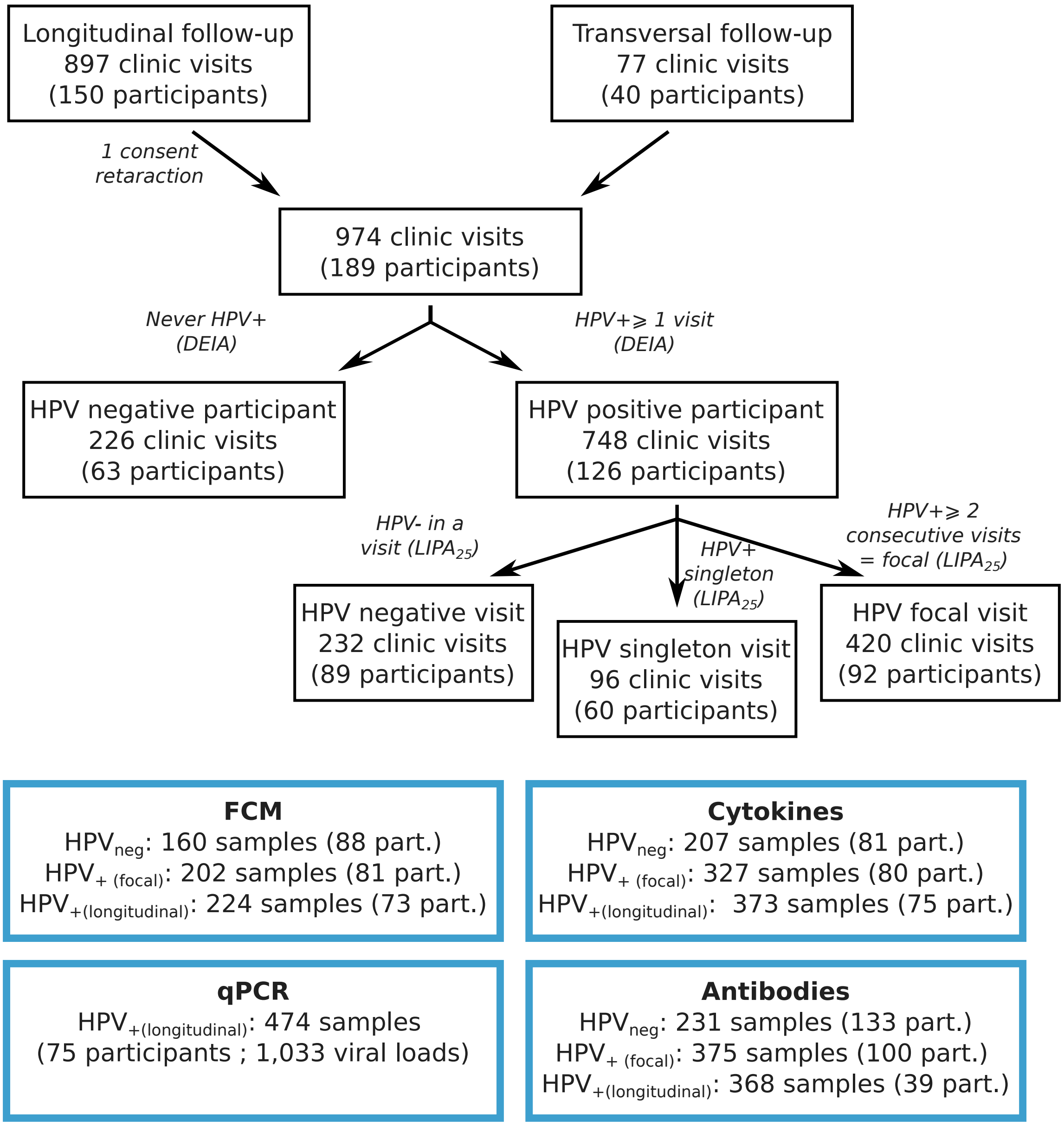

Supplementary Figure S1: **Diagram of the sample selection.** The bottom boxes indicate the number and nature of samples used in each type of analysis.

#### S2.4 Flow cytometry clustering analyses

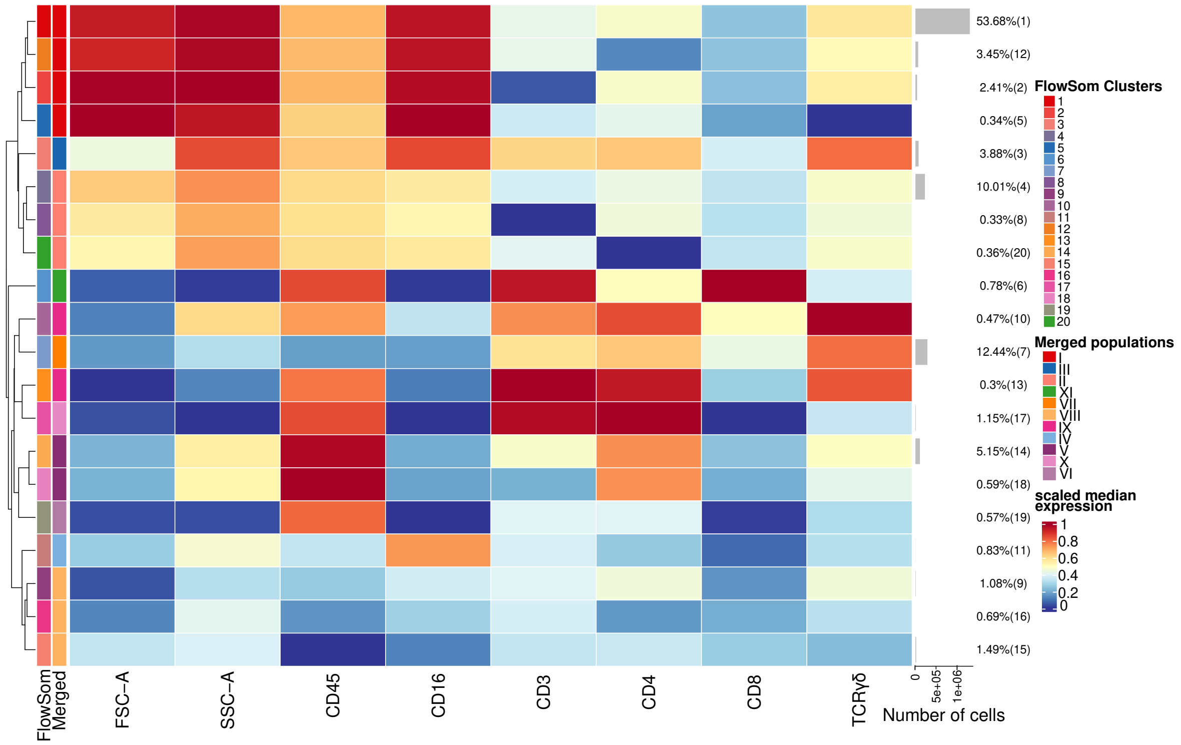

Supplementary Figure S2: **Heatmap of 20 FlowSom metaclusters and their annotation in 11 immune cells populations.** Twenty FlowSom metaclusters are identified on the left. Their merging and annotation into 11 immune cells populations is indicated in the second column names 'Merged'. Fluorescence is scaled on a per marker basis. The total number of cells is shown in grey bar charts. The scale for the heatmap, which shows fluorescence intensity, is shown on the right.

| Cluster | Markers | Identification | Commentary |
|---|---|---|---|
| I | $FSC^{high}$ $SSC^{high}$ $CD45^{int}$ $CD3^{-}$ $CD4^{-}$ $CD16^{+}$ $TCR\gamma\delta^{-}$ | $FSC^{high}$ $SSC^{high}$ $CD16^{high}$ cells | Probably majority of neutrophils |
| II | $FSC^{int}$ $SSC^{int}$ $CD16^{+}$ cells | $FSC^{int}$ $SSC^{int}$ $CD16^{+}$ cells | Possible NK cells, macrophages or Langerhans |
| III | $FSC^{int}$ $SSC^{hi}$ $CD45^{int}$ $CD3^{lo}$ $CD4^{lo}$ $CD16^{-}$ $TCR\gamma\delta^{int}$ | $CD16^{+}$ $TCR\gamma\delta$ cells | |
| IV | $FSC^{lo}$ $SSC^{int}$ $CD45^{int}$ $CD3^{-}$ $CD4^{-}$ $CD16^{int}$ $TCR\gamma\delta^{-}$ | $CD45^{low}$ $CD16^{+}$ leukocytes | Possible NK cells |
| V | $FSC^{lo}$ $SSC^{int}$ $CD45^{+}$ $CD3^{-}$ $CD4^{int}$ $CD16^{-}$ $TCR\gamma\delta^{-}$ | $CD45^{+}$ $CD4^{int}$ leukocytes | Possible Monocytes, Macrophages or Langerhans cells |
| VI | $FSC^{lo}$ $SSC^{lo}$ $CD45^{+}$ $CD3^{-}$ $CD4^{-}$ $CD16^{-}$ $TCR\gamma\delta^{-}$ | $CD45^{+}$ $CD3^{-}$ $CD16^{-}$ leukocytes | Possible B cells or ILCs |
| VII | $FSC^{lo}$ $SSC^{int}$ $CD45^{lo}$ $CD3^{lo}$ $CD4^{lo}$ $CD16^{-}$ $TCR\gamma\delta^{int}$ | $CD45^{low}$ $TCR\gamma\delta$ cells | |
| VIII | $FSC^{lo}$ $SSC^{int}$ $CD45^{lo}$ $CD3^{-}$ $CD4^{-}$ $CD16^{-}$ $TCR\gamma\delta^{-}$ | $CD45^{low}$ $CD3^{-}$ $CD16^{-}$ cells | Possible B cells, ILCs or progenitors |
| IX | $FSC^{lo}$ $SSC^{lo}$ $CD45^{+}$ $CD3^{int}$ $CD4^{+}$ $CD16^{-}$ $TCR\gamma\delta^{+}$ | $CD45^{+}$ $TCR\gamma\delta$ cells | |
| X | $FSC^{lo}$ $SSC^{lo}$ $CD45^{+}$ $CD3^{+}$ $CD4^{+}$ | CD4 T cells | |
| XI | $FSC^{lo}$ $SSC^{lo}$ $CD45^{+}$ $CD3^{+}$ $CD8^{+}$ | CD8 T cells | Possibly includes MAIT cells |

Table S3: **Immune cell clusters identification and annotation.** 'Markers' indicates the fluorescence intensity of the key flow cytometry labels used to annotate each clusters and 'Commentary' indicates possible alternatives or characteristics of each cluster.

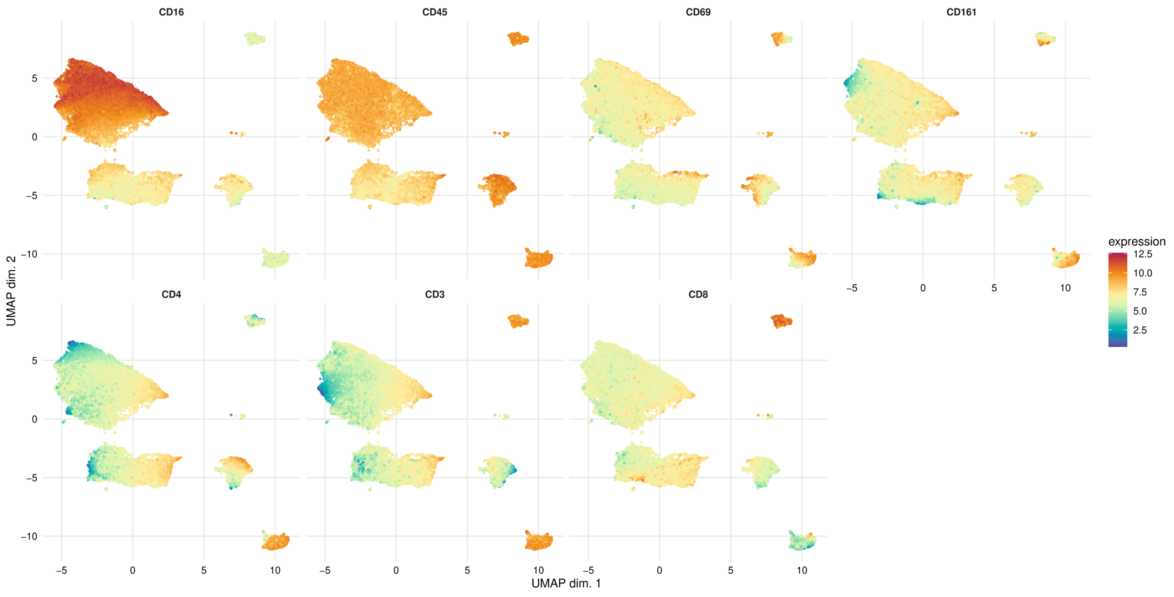

Supplementary Figure S3: **Individual UMAPs per flow cytometry fluorescent label used in the FlowSom clustering.** Scaled intensity of each fluorescent marker is individually represented on the global UMAP displayed in Fig. 1A

.

| Cluster | Marker | FC | p_val | p_adj |
|---|---|---|---|---|
| I | CD69 | 1.1610 | 0.0000 | 0.0000 |
| II | CD69 | 1.1504 | 0.0000 | 0.0000 |
| III | CD69 | 1.1916 | 0.0000 | 0.0000 |
| IV | CD69 | 1.0820 | 0.0227 | 0.0340 |
| V | CD69 | 1.2996 | 0.0000 | 0.0000 |
| VI | CD69 | 0.9396 | 0.2407 | 0.2941 |
| VII | CD69 | 1.0317 | 0.2887 | 0.3285 |
| VIII | CD69 | 1.2023 | 0.0000 | 0.0000 |
| IX | CD69 | 1.3375 | 0.0000 | 0.0000 |
| X | CD69 | 0.8991 | 0.0278 | 0.0383 |
| XI | CD69 | 1.0607 | 0.3458 | 0.3804 |
| I | CD161 | 1.2788 | 0.0000 | 0.0000 |
| II | CD161 | 1.2544 | 0.0000 | 0.0000 |
| III | CD161 | 1.1602 | 0.0000 | 0.0000 |
| IV | CD161 | 1.3329 | 0.0000 | 0.0000 |
| V | CD161 | 1.3559 | 0.0000 | 0.0000 |
| VII | CD161 | 1.0985 | 0.0032 | 0.0050 |
| VI | CD161 | 1.0962 | 0.0260 | 0.0373 |
| VIII | CD161 | 0.7944 | 0.0005 | 0.0008 |
| IX | CD161 | 1.5443 | 0.0000 | 0.0000 |
| X | CD161 | 0.9648 | 0.4044 | 0.4305 |
| XI | CD161 | 0.9079 | 0.0618 | 0.0816 |

Table S4: **Differential expression analysis.** Fold change (FC) for CD69 and CD161 fluorescence intensity for each of the 11 immune cells populations identified in flow cytometry analyses. Fold change value of HPV positive (focal) over HPV negative and the adjusted p-value. Fold changes were calculated by an abundance analysis performed with diffcyt-DS-limma and adjusted with a Benjamini-Hochberg test.

#### S2.5 Local cytokine concentrations

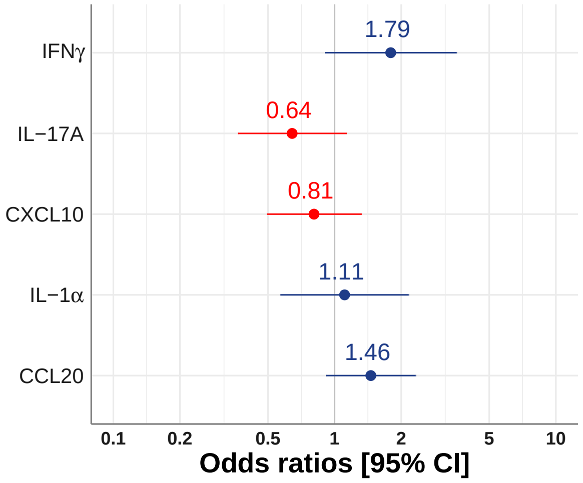

Supplementary Figure S4: **Cytokines and chemokines concentrations in HPV focal infections vs. negative samples.** Odds ratios were calculated with HPV positive (focal) as a response, the concentration of the five cytokines and chemokines as covariates, and a random effect for the participant.

#### S2.6 Viral kinetics

##### S2.6.1 Parameters estimates

Table S5: **Parameters estimates for the viral kinetics three-slope model.** We summarise the posterior distribution of each parameter, which is an aggregation of the 3,000 samplings on 4 MCMC chains, with its mean value, standard deviation, $2.5^{th}$, $50^{th}$ and $97.5^{th}$ quantiles. $\hat{R}$ is a metric indicating if the chains have converged (and then tends to 1), and $N_{\text{eff}}$ is the effective sample size, correcting for the potential autocorrelation between the samples within a chain. $\mu$ represent the fixed effects, $\omega$ the diagonal parameters of the variance-covariance matrix of the random effects, and $\sigma_{\text{vl}}$ is the additive measurement error.

| parameter | mean | sd | 2.5% | 50% | 97.5% | n_eff | Rhat |
|---|---|---|---|---|---|---|---|
| $\mu_a$ | 1.16 | 0.33 | 0.54 | 1.15 | 1.84 | 5607 | 1.00 |
| $\mu_b$ | 12.33 | 1.35 | 9.80 | 12.28 | 15.10 | 3776 | 1.00 |
| $\mu_c$ | 0.92 | 0.26 | 0.43 | 0.91 | 1.46 | 5868 | 1.00 |
| $\mu_d$ | 5.03 | 0.16 | 4.72 | 5.03 | 5.33 | 6728 | 1.00 |
| $\mu_{mid}$ | 4.68 | 0.85 | 3.02 | 4.70 | 6.31 | 4072 | 1.00 |
| $\omega_1$[1] | 0.68 | 0.17 | 0.41 | 0.67 | 1.05 | 1645 | 1.01 |
| $\omega_1$[2] | 0.17 | 0.03 | 0.12 | 0.17 | 0.24 | 3549 | 1.00 |
| $\omega_1$[3] | 6.37 | 0.71 | 5.09 | 6.33 | 7.87 | 3886 | 1.00 |
| $\omega_2$[1] | 1.32 | 0.18 | 1.00 | 1.31 | 1.72 | 3665 | 1.00 |
| $\omega_2$[2] | 0.25 | 0.03 | 0.20 | 0.25 | 0.31 | 4279 | 1.00 |
| $\omega_2$[3] | 6.29 | 0.64 | 5.15 | 6.25 | 7.65 | 5605 | 1.00 |
| $\sigma_{vl}$ | 1.15 | 0.05 | 1.06 | 1.15 | 1.25 | 1700 | 1.00 |

##### S2.6.2 Individual infection durations

At the infection level, the median of the infection duration posterior distribution ranged from 1.6 to 40.2 months. Therefore, our maximal plateau duration of 60 months was only reached on rare occasions for some infections where both the beginning and the end were censored. As expected, complete follow-ups were on average shorter than follow-ups where the beginning or the end was missing, which themselves were shorter than follow-ups that missed both the begining and end.

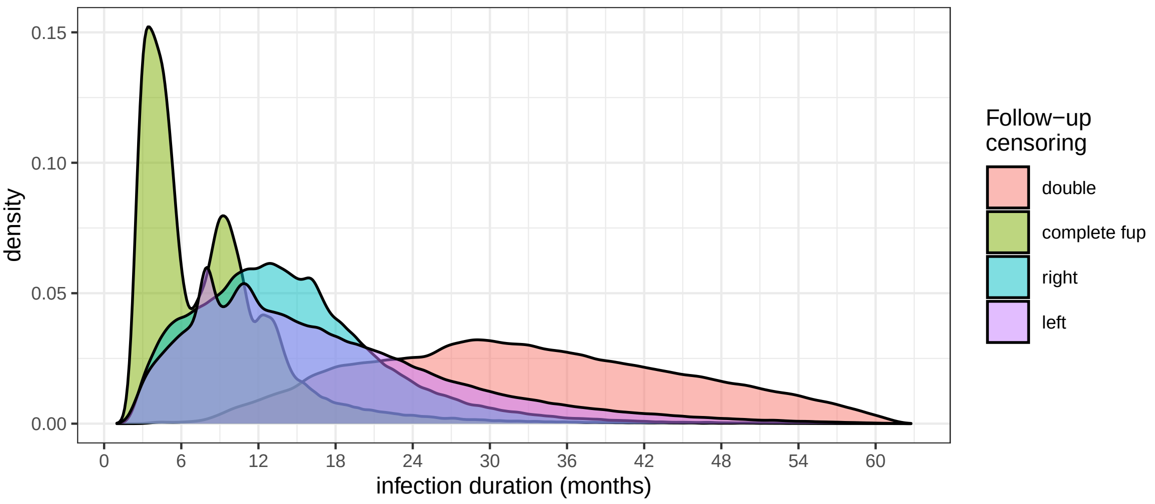

Supplementary Figure S5: **Posterior distribution of the infections durations.** This density plot represents the infection duration of all participants and all MCMC samples. Colors indicate the type of follow-up censoring (if any).

##### S2.6.3 Correlation between plateau level and infection duration

Our model included a correlation parameter between the random effects of $\psi_b$ and $\psi_d$. The correlation between the two parameters was not significantly higher than zero. It was small at the between-host level (0.080, 95% CrI -0.44; 0.53) and higher at the within-host level (0.21, 95% CrI -0.10; 0.50).

However, we could observe a significant correlation of 0.29 ($p = 4.10^{-4}$) between the median infection duration and the median viral load peak at plateau.

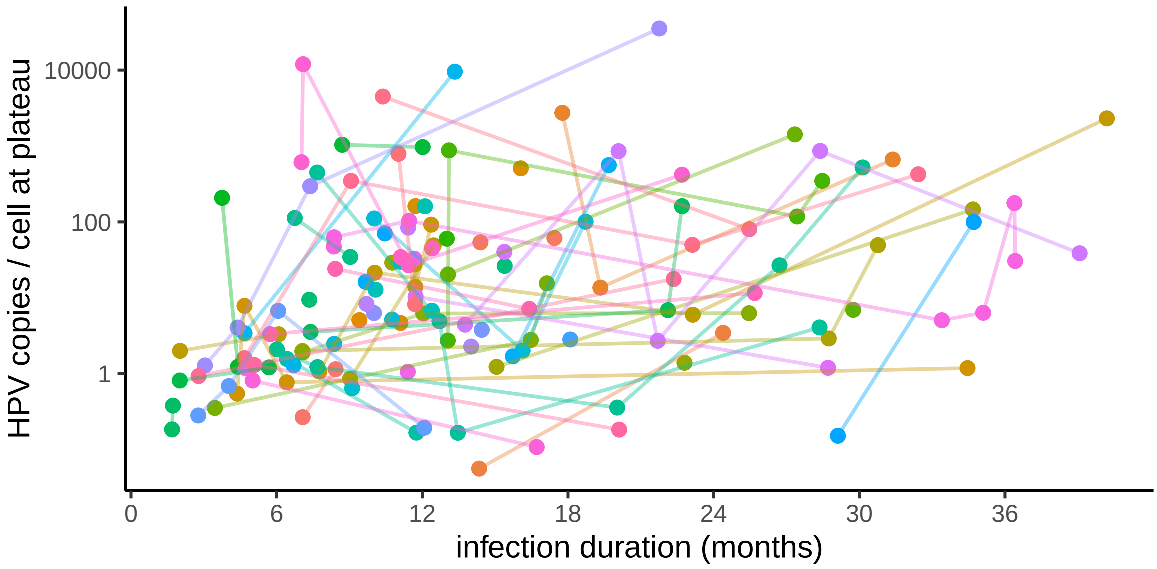

Supplementary Figure S6: **Plateau viral load as a function of the posterior median infection duration.** Each dot represents one infection, and each color represents an individual, with the lines connecting the infections of a same individual. The between-host correlation between those two parameters is very low (posterior median: 0.08, 95% CrI: -0.44 ; 0.52), but there is a trend towards as stronger correlation at the within-host level (posterior median: 0.21, 95% CrI: -0.10 ; 0.50)

#### S2.7 Immune response kinetics modelling

##### S2.7.1 Parameters estimates

Table S6: **Parameters estimates for the cytokines two-slope model.** We summarise the posterior distribution of each parameter as Table S5. $\mu$s represent the fixed effects for each parameter $\psi_e$, $\psi_f$, $\psi_g$ and each cytokine, as well as for $\sigma_{\text{cyt}}$, which is the additive measurement error.

| parameter | mean | sd | 2.5% | 50% | 97.5% | n_eff | Rhat |
|---|---|---|---|---|---|---|---|
| $\mu_{e_{\text{IFN}\gamma}}$ | 1.01 | 0.08 | 0.85 | 1.01 | 1.18 | 1710 | 1.00 |
| $\mu_{f_{\text{IFN}\gamma}}$ | -0.18 | 0.14 | -0.45 | -0.18 | 0.08 | 3200 | 1.00 |
| $\mu_{g_{\text{IFN}\gamma}}$ | 0.18 | 0.15 | 0.02 | 0.14 | 0.58 | 1490 | 1.00 |
| $\mu_{e_{\text{IL17a}}}$ | 0.87 | 0.10 | 0.69 | 0.87 | 1.06 | 1910 | 1.00 |
| $\mu_{f_{\text{IL17a}}}$ | -0.08 | 0.15 | -0.38 | -0.08 | 0.21 | 4320 | 1.00 |
| $\mu_{g_{\text{IL17a}}}$ | 0.20 | 0.10 | 0.04 | 0.19 | 0.41 | 2410 | 1.00 |
| $\mu_{e_{\text{CXCL10}}}$ | 2.77 | 0.10 | 2.56 | 2.77 | 2.98 | 470 | 1.01 |
| $\mu_{f_{\text{CXCL10}}}$ | -0.01 | 0.18 | -0.36 | -0.00 | 0.34 | 296 | 1.02 |
| $\mu_{g_{\text{CXCL10}}}$ | 0.28 | 0.17 | 0.04 | 0.25 | 0.69 | 134 | 1.03 |
| $\mu_{e_{\text{CCL20}}}$ | 1.95 | 0.09 | 1.78 | 1.95 | 2.13 | 1450 | 1.01 |
| $\mu_{f_{\text{CCL20}}}$ | 0.15 | 0.14 | -0.12 | 0.15 | 0.43 | 1130 | 1.01 |
| $\mu_{g_{\text{CCL20}}}$ | 0.40 | 0.15 | 0.14 | 0.39 | 0.71 | 229 | 1.02 |
| $\mu_{e_{\text{IL1}\alpha}}$ | 3.15 | 0.08 | 2.99 | 3.15 | 3.31 | 2670 | 1.00 |
| $\mu_{f_{\text{IL1}\alpha}}$ | -0.05 | 0.13 | -0.30 | -0.05 | 0.19 | 2910 | 1.00 |
| $\mu_{g_{\text{IL1}\alpha}}$ | 0.43 | 0.26 | 0.04 | 0.39 | 0.93 | 2290 | 1.00 |
| $\sigma_{\text{cyt}}$ | 0.51 | 0.00 | 0.49 | 0.51 | 0.54 | 3000 | 1.00 |

Table S7: **Parameters estimates for the FCM two-slope model.** We summarise the posterior distribution of each parameter as in Table S5. The fixed effects parameters ($\mu$s) are stored in a matrix with each row representing one cell cluster, and each column the parameter in the following order: $\mu_e$, $\mu_f$, $\mu_g$, and $\sigma_{\text{FCM}}$, which is the additive measurement error.

| | mean | sd | 2.5% | 50% | 97.5% | n_eff | Rhat |
|---|---|---|---|---|---|---|---|
| mu[1,1] | 1.28 | 0.22 | 0.85 | 1.28 | 1.72 | 3260.00 | 1.00 |
| mu[1,2] | 0.83 | 0.43 | -0.03 | 0.84 | 1.68 | 3850.00 | 1.00 |
| mu[1,3] | 0.22 | 0.10 | 0.05 | 0.22 | 0.43 | 2900.00 | 1.00 |
| mu[2,1] | 0.98 | 0.20 | 0.59 | 0.98 | 1.40 | 3440.00 | 1.00 |
| mu[2,2] | 0.65 | 0.39 | -0.15 | 0.66 | 1.38 | 3370.00 | 1.00 |
| mu[2,3] | 0.26 | 0.19 | 0.03 | 0.22 | 0.78 | 2160.00 | 1.00 |
| mu[3,1] | -0.79 | 0.20 | -1.18 | -0.79 | -0.39 | 3720.00 | 1.00 |
| mu[3,2] | -0.31 | 0.39 | -1.08 | -0.30 | 0.46 | 4030.00 | 1.00 |
| mu[3,3] | 0.34 | 0.21 | 0.03 | 0.32 | 0.80 | 2170.00 | 1.00 |
| mu[4,1] | -1.20 | 0.20 | -1.60 | -1.20 | -0.81 | 4670.00 | 1.00 |
| mu[4,2] | -0.11 | 0.36 | -0.80 | -0.11 | 0.60 | 4720.00 | 1.00 |
| mu[4,3] | 0.45 | 0.24 | 0.05 | 0.43 | 0.92 | 3240.00 | 1.00 |
| mu[5,1] | 0.53 | 0.17 | 0.18 | 0.53 | 0.87 | 4720.00 | 1.00 |
| mu[5,2] | -0.59 | 0.38 | -1.36 | -0.59 | 0.12 | 3980.00 | 1.00 |
| mu[5,3] | 0.25 | 0.20 | 0.02 | 0.20 | 0.77 | 1760.00 | 1.00 |
| mu[6,1] | -1.30 | 0.19 | -1.67 | -1.30 | -0.91 | 4970.00 | 1.00 |
| mu[6,2] | -0.18 | 0.35 | -0.85 | -0.18 | 0.51 | 5200.00 | 1.00 |
| mu[6,3] | 0.51 | 0.25 | 0.07 | 0.51 | 0.94 | 3820.00 | 1.00 |
| mu[7,1] | 0.96 | 0.20 | 0.57 | 0.96 | 1.33 | 3560.00 | 1.00 |
| mu[7,2] | 0.16 | 0.39 | -0.59 | 0.16 | 0.92 | 3410.00 | 1.00 |
| mu[7,3] | 0.41 | 0.23 | 0.05 | 0.40 | 0.85 | 1820.00 | 1.00 |
| mu[8,1] | 0.93 | 0.23 | 0.48 | 0.93 | 1.37 | 3010.00 | 1.00 |
| mu[8,2] | 0.40 | 0.42 | -0.41 | 0.40 | 1.24 | 3560.00 | 1.00 |
| mu[8,3] | 0.27 | 0.21 | 0.02 | 0.20 | 0.79 | 966.00 | 1.00 |
| mu[9,1] | -2.22 | 0.19 | -2.59 | -2.21 | -1.83 | 3780.00 | 1.00 |
| mu[9,2] | -0.30 | 0.39 | -1.08 | -0.29 | 0.46 | 3480.00 | 1.00 |
| mu[9,3] | 0.44 | 0.24 | 0.05 | 0.42 | 0.92 | 2710.00 | 1.00 |
| mu[10,1] | -0.38 | 0.22 | -0.83 | -0.38 | 0.04 | 3680.00 | 1.00 |
| mu[10,2] | -0.82 | 0.42 | -1.60 | -0.83 | 0.07 | 3680.00 | 1.00 |
| mu[10,3] | 0.25 | 0.17 | 0.03 | 0.22 | 0.69 | 2370.00 | 1.00 |
| mu[11,1] | -0.33 | 0.20 | -0.74 | -0.33 | 0.04 | 3270.00 | 1.00 |
| mu[11,2] | -0.46 | 0.40 | -1.21 | -0.47 | 0.36 | 3140.00 | 1.00 |
| mu[11,3] | 0.34 | 0.21 | 0.04 | 0.29 | 0.85 | 2140.00 | 1.00 |
| $\sigma_{\text{FCM}}$ | 1.38 | 0.03 | 1.33 | 1.38 | 1.44 | 1980.00 | 1.00 |

Table S8: **Parameters estimates for the IgG Bateman model.** We summarise the posterior distribution of each parameter as in Table S5. $\mu$ represent the fixed effects, $\omega$ the random effect standard deviation, $\sigma_1$ the additive measurement error, and $\sigma_2$ the proportional measurement error.

| | mean | sd | 2.5% | 50% | 97.5% | n_eff | Rhat |
|---|---|---|---|---|---|---|---|
| $\mu_h$ | 6.04 | 1.89 | 2.68 | 5.92 | 10.20 | 3408.12 | 1.00 |
| $\mu_k$ | 48.78 | 10.12 | 30.90 | 48.14 | 70.40 | 6101.39 | 1.00 |
| $\mu_l$ | 8.29 | 0.18 | 7.94 | 8.29 | 8.66 | 1666.52 | 1.00 |
| $\mu_m$ | 25.21 | 2.87 | 20.39 | 24.94 | 31.64 | 1565.98 | 1.00 |
| $\omega_4$[1] | 1.35 | 0.40 | 0.68 | 1.32 | 2.26 | 2522.68 | 1.00 |
| $\omega_4$[2] | 0.51 | 0.37 | 0.03 | 0.45 | 1.36 | 819.44 | 1.00 |
| $\omega_4$[3] | 0.76 | 0.12 | 0.55 | 0.75 | 1.02 | 3310.83 | 1.00 |
| $\omega_4$[4] | 0.45 | 0.11 | 0.27 | 0.44 | 0.71 | 811.46 | 1.00 |
| $\sigma_1$ | 26.45 | 21.58 | 0.90 | 21.18 | 80.42 | 6557.84 | 1.00 |
| $\sigma_2$ | 0.26 | 0.02 | 0.22 | 0.26 | 0.31 | 1681.01 | 1.00 |

Table S9: **Parameters estimates for the IgM Bateman model.** We summarise the posterior distribution of each parameter as in Table S5. $\mu$ represent the fixed effects, $\omega$ the random effect standard deviation, $\sigma_1$ the additive measurement error, and $\sigma_2$ the proportional measurement error.

| | mean | sd | 2.5% | 50% | 97.5% | n_eff | Rhat |
|---|---|---|---|---|---|---|---|
| $\mu_h$ | 1.64 | 1.14 | 0.04 | 1.48 | 4.29 | 217.21 | 1.02 |
| $\mu_k$ | 21.86 | 5.78 | 12.37 | 21.22 | 34.65 | 248.80 | 1.01 |
| $\mu_l$ | 7.43 | 0.29 | 6.91 | 7.41 | 8.02 | 145.21 | 1.01 |
| $\mu_m$ | 22.73 | 2.91 | 17.55 | 22.58 | 29.17 | 184.88 | 1.01 |
| $\omega_4$[1] | 2.38 | 0.93 | 0.66 | 2.35 | 4.32 | 140.44 | 1.03 |
| $\omega_4$[2] | 1.05 | 0.56 | 0.32 | 0.93 | 2.73 | 57.31 | 1.06 |
| $\omega_4$[3] | 0.69 | 0.19 | 0.35 | 0.67 | 1.11 | 479.87 | 1.00 |
| $\omega_4$[4] | 0.52 | 0.24 | 0.25 | 0.45 | 1.25 | 57.03 | 1.07 |
| $\sigma_1$ | 279.06 | 36.92 | 210.92 | 278.47 | 357.84 | 181.65 | 1.02 |
| $\sigma_2$ | 0.03 | 0.01 | 0.00 | 0.02 | 0.06 | 378.00 | 1.01 |

##### S2.7.2 Random effects correlation matrix

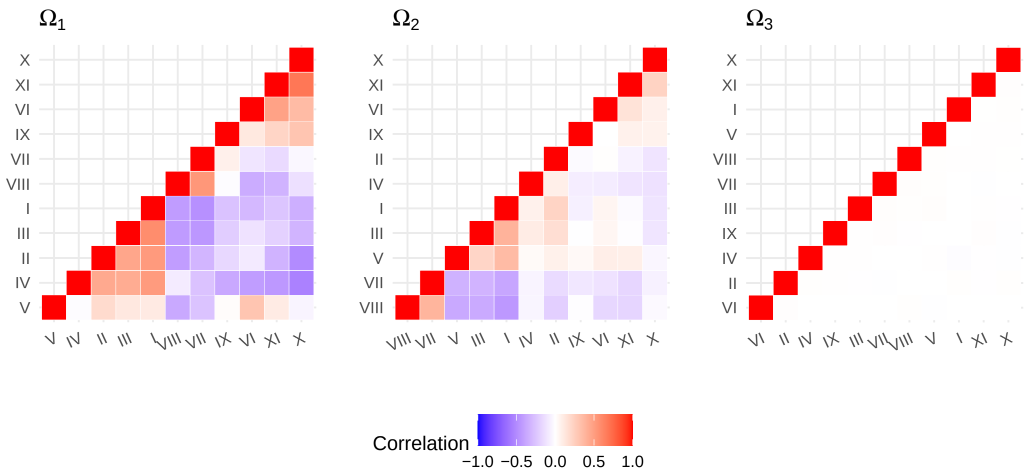

Supplementary Figure S7: **Correlation between random effects of the different cell clusters.** Each parameter $\psi_e$, $\psi_f$, and $\psi_g$ is associated with a variance-covariance matrix for the random effects between the cell clusters (respectively $\Omega_1$, $\Omega_2$, and $\Omega_3$). Notice that the covariance of the random effect associated with parameter $\psi_3$ (governing the peak timing), $\Omega_3$, is close to zero.

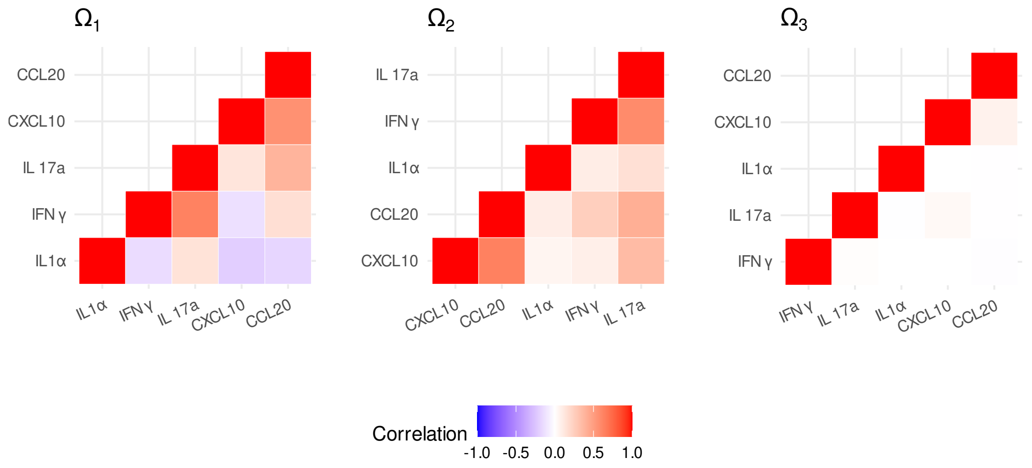

Supplementary Figure S8: **Correlation between random effects of the different cytokines.** Each parameter $\psi_e$, $\psi_f$, and $\psi_g$ is associated with a variance-covariance matrix for the random effects between the cytokines, respectively $\Omega_1$, $\Omega_2$ and $\Omega_3$. The covariance of the random effect associated with parameter $\psi_3$ (governing the peak timing), $\Omega_3$, is close to zero.

##### S2.7.3 Cells populations associated with the IgG serum titer

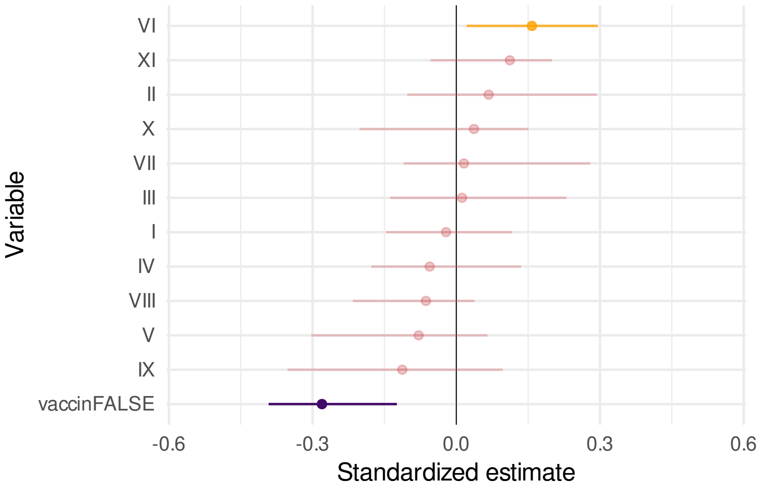

Supplementary Figure S9: **PLSR regression of the IgG mean serum titer as a function of the mean cells clusters frequencies and vaccine status.** Not being vaccinated is associated with a significantly lower IgG serum titer, regardless of the HPV genotype, which indicates a potential cross-protection. Cell cluster VI mean frequency (log-centered) is significantly correlated with the titer of IgGs specific to the HPV genotype causing the infection.

### S3 Ethics, competing interests, and authors' contribution

#### S3.1 Ethics

This study has been approved by the Comité de Protection des Personnes (CPP) Sud Méditerranée I (reference number 2016-A00712-49); by the Comité Consultatif sur le Traitement de l'Information en matière de Recherche dans le domaine de la Santé (reference number 16.504); by the Commission Nationale Informatique et Libertés (reference number MMS/ABD/AR1612278, decision number DR-2016–488), by the Agence Nationale de Sécurité du Médicament et des Produits de Santé (reference 20160072000007), and is registered at ClinicalTrials.gov under the ID NCT02946346.

#### S3.2 Competing interests

T.W. serves on advisory boards for MSD (Merck Sharp and Dohme).

J. R. reports personal fees from Gilead (consulting and payment or honoraria for lectures, presentations, speaker's bureaus, manuscript writing, or educational events), Janssen (payment or honoraria for lectures, presentations, speaker's bureaus, manuscript writing, or educational events), Merck (payment or honoraria for lectures, presentations, speaker's bureaus, manuscript writing, or educational events), Theratechnologies (payment or honoraria for lectures, presentations, speaker's bureaus, manuscript writing, or educational events), and ViiV Healthcare (consulting and payment or honoraria for lectures, presentations, speaker's bureaus, manuscript writing, or educational events) and support for attending meetings and/or travel from Gilead and Pfizer, outside of the submitted work.

All the other authors do not report any conflict of interest.

#### S3.3 Authors contribution

Nicolas Tessandier structured and analysed the databases, implemented the flow cytometry data, and performed all the immunological analyses.

Baptiste Elie structured and analysed the databases, and performed the Bayesian hierarchical modelling.

Christian Selinger structured and analysed the databases.

Claire Bernat implemented some of the laboratory protocols and processed some of the samples.

Vanina Boué implemented the qPCR protocols and processed some of the samples, including all the qPCR experiments.

Soraya Groc processed some of the samples.

Massilva Rahmoun provided immunological and flow cytometry expertise and coordinated the clinical study and the sample processing.

Bastien Reyné helped to structure and analyse the databases.

Anne-Sophie Bedin provided expertise for the flow cytometry protocol and analyses.

Thomas Beneteau provided statistical expertise.

Marine Bonneau performed some of the clinical visits and provided expertise.

Christelle Graf performed some of the clinical visits and provided expertise.

Jérémie Guedj provided critical statistical expertise.

Nathalie Jacobs provided critical immunological and flow cytometry expertise.

Tsukushi Kamiya provided critical statistical expertise.

Marion Kerioui provided critical statistical expertise.

Julie Lajoie provided critical immunological and flow cytometry expertise.

Imène Melki provided critical immunological and flow cytometry expertise.

Jean-Luc Prétet provided virology expertise to set up the qPCR.

Géraldine Schlecht-Louf provided critical immunological expertise and helped with the interpretation of the flow cytometry data.

Mircea T. Sofonea provided critical statistical expertise.

Tim Waterboer provided critical immunological expertise and his team performed the antibody titrations.

Christophe Hirtz provided immunological expertise and his team performed the cytokines dosages.

Vincent Foulongne provided virology expertise and his laboratory hosted most of the experiments.

Marie-Christine Picot provided epidemiological expertise and her team set up the case reporting form.

Jacques Reynes provided epidemiological expertise and was the lead clinical investigator in the study.

Vincent Tribout provided epidemiological expertise and his team of the STI detection centre performed the clinical visits.

Édouard Tuaillon provided immunological expertise and his laboratory provided the support for the flow cytometry analyses.

Ignacio G Bravo provided critical expertise on HPVs and in virology in general.

Michel Segondy was one of the co-PIs of the clinical study, provided virology expertise and his laboratory hosted most of the experiments.

Nathalie Boulle was one of the co-PIs of the clinical study, provided HPV expertise and her laboratory performed cervical lesion screening.

Carmen Lia Murall contributed to the conception of the project, the implementation of the clinical study, and to the data analysis.

Samuel Alizon conceived the project, implemented the clinical study, structured and analysed the databases, and wrote the first version of the manuscript.

S17. A. Canty, B. D. Ripley, *boot: Bootstrap R (S-Plus) Functions*, R package version 1.3-28.1.
